# A Trans-Omic Mendelian Randomization Study of Parental Lifespan Uncovers Novel Aging Biology and Drug Candidates for Human Healthspan Extension

**DOI:** 10.1101/2020.07.06.20147520

**Authors:** Nicolas Perrot, William Pelletier, Jérôme Bourgault, Christian Couture, Zhonglin Li, Patricia Mitchell, Nooshin Ghodsian, Yohan Bossé, Sébastien Thériault, Patrick Mathieu, Benoit J. Arsenault

**Author notes:** Address for correspondence Benoit Arsenault, PhD, Centre de recherche de l’Institut universitaire de cardiologie et de pneumologie de Québec – Université Laval, Y-3106, Pavillon Marguerite D’Youville, 2725 chemin Ste-Foy, Québec (QC) Canada G1V 4G5.

## Abstract

The study of parental lifespan has emerged as an innovative tool to advance aging biology and our understanding of the genetic architecture of human longevity and healthspan. Here, we leveraged summary statistics of a genome-wide association study including over one million parental lifespans to identify genetically regulated genes from the Genotype-Tissue Expression project through a combination of multi-tissue transcriptome-wide association analyses and genetic colocalization. Mendelian randomization (MR) analyses also identified circulating proteins and metabolites causally associated with parental lifespan that may offer new drug repositioning opportunities for healthspan such as drugs targeting apolipoprotein-B-containing lipoproteins. Liver expression of *HP*, the gene encoding haptoglobin, and plasma haptoglobin levels were causally linked with parental lifespan. Phenome-wide MR analyses were used to map genetically regulated genes, proteins and metabolites with the disease-related phenome in the UK Biobank and FinnGen. Altogether, this study identified novel biological determinants of aging and potential therapeutic targets for human healthspan extension.

## INTRODUCTION

The genetics revolution offers new opportunities to foster our understanding of complex traits such as human longevity^1,2^. The definition of longevity in human genetic studies is highly debated and the lack of a universally recognized definition increases the possibility of biases and limits external validation. Studying the genetic architecture of long-lived individuals has shed some light on the loci at which genetic variation might influence human longevity. However, the most comprehensive meta-analysis of genome-wide association studies (GWAS) of survival percentiles only reported one locus (*APOE*) to be robustly associated with human longevity^3^. On the other hand, other longevity traits such as parental lifespan (the lifespan of deceased or long-lived parents) have also been used to uncover longevity genes. In the most comprehensive GWAS of parental lifespan published to date, Timmers et al.^4^ reported several new loci associated with parental lifespan, including *APOE*. Although it has been suggested that human longevity is not simply limited to the absence life-threatening diseases, many of the reported genetic variants associated with lifespan are also associated with chronic diseases. A better characterization of longevity genomics could therefore contribute to the identification of drug development strategies to enhance “healthspan”, i.e. the number of years of living without chronic diseases.

Mendelian randomization (MR) is a burgeoning field of research that draws on the use of genetic variants as instruments to assess potentially causal relationships between a wide variety of exposures such as risk factors or drug targets and outcomes such as age-related disease or human longevity^5^. By taking advantage of the naturally randomized allocation of genetic variation MR is not subject to many of the biases of observational studies such as reverse causality or random measurement error and less susceptible to confounding and reverse causality. The recent public release of a wide-variety of deeply phenotyped gene-trait association datasets has enabled the use of multi-omic datasets as exposures in MR studies. Few studies however have investigated the long-term consequences of lifelong exposure to over- or under-expressed genes (eGenes), plasma proteins, lipoproteins or metabolites on human lifespan and associated age-related chronic disease. In this study, we aimed at discovering and harnessing novel biological determinants of human longevity by identifying eGenes through transcriptome-wide association study (TWAS) as well as genetically regulated circulating proteins (eProteins) and metabolites (eMetabolites) using a highly translational MR framework (Fig. 1). We performed a TWAS for parental lifespan leveraging expression Quantitative Trait Loci (eQTL) data from 43 non-sex-specific tissues to identify non-pleiotropic and colocalized eGenes associated with over one million parental lifespans. We also used MR to identify key eProteins and eMetabolites that may regulate human lifespan. In order to discover the role of these genetically-regulated traits in human homeostasis while at the same time determining whether they could be effectively and safely targeted to promote healthy aging and human longevity, we report MR findings across the human disease-related phenome.

**Figure 1.**
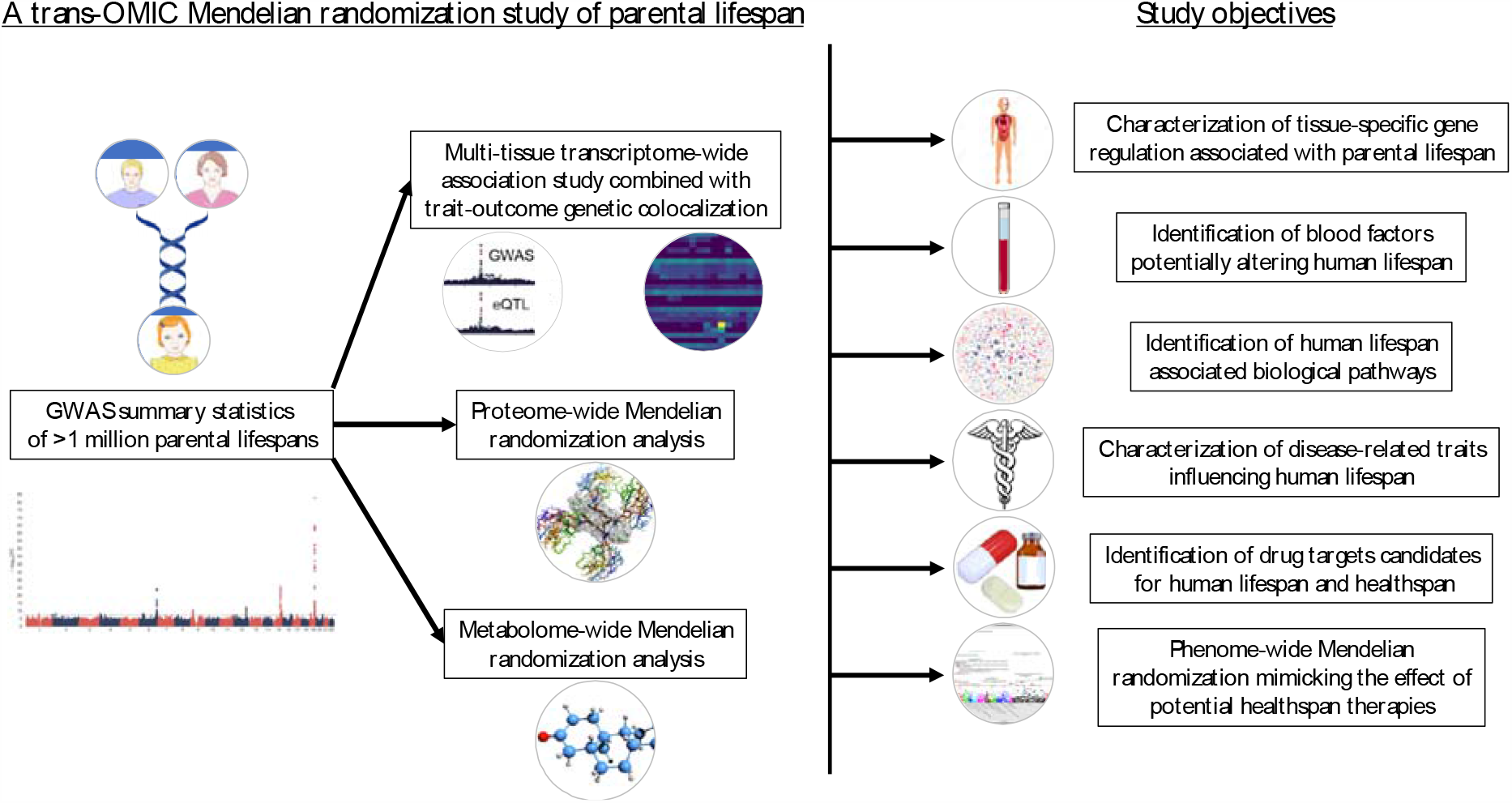
Schematic overview of the analytical framework used to identify novel biological determinants and potential drug targets for healthspan using genome-wide association study summary statistics of >1 million parental lifespans.

## RESULTS

### Identification of eGenes associated with parental lifespan

We performed a TWAS using the MetaXcan framework^6^ to identify potentially causal effects of eGenes on parental lifespan using GWAS summary-level statistics of the UK Biobank and LifeGen consortium meta-analysis (methods)^4^. This analytical approach uses eQTL mapping from the Genotype-Tissue Expression (GTEx) project across 43 non-sex-specific tissues to estimate gene-level association with summary-level GWAS results. We identified several new potential eGenes for parental lifespan after correction for multiple testing (Fig 2a). A detailed description of the association of eGenes-parental lifespan associations across all tissues is presented in Supplementary Table 1. Because TWAS prioritizes multiple genes, some of the eGenes could be non-causal, owing to sharing of eQTLs or co-regulated genes. TWAS may thus be prone to false positives and spurious gene prioritization, especially in the absence of genetic colocalization, that is, when the lead genetic variant driving gene expression is not amongst the lead GWAS variants. We therefore took additional steps using genetic colocalization to control for spurious prioritization. After excluding genes with a posterior probability of statistical colocalization <0.75 and after excluding genes found in pleiotropic regions such as *HLA, ABO* and *APOE*, the number of parental lifespan eGenes was reduced to 30, which span 17 loci. Details on strength of the association, colocalization of these eGenes with parental lifespan as well as the “lead tissue” (the tissue that provided the strongest eGenes-parental lifespan estimate from MetaXcan) are provided in Table 1. This analysis identified tissue-specific expression regulation of several known parental lifespan genes and also revealed potentially new parental lifespan genetic signals at the *LRP8* (ApoER2), *NEK10, CCDC71L, NRG1* and *RAD52* loci. Although the use of statistical colocalization helped prioritize several genes, the causal gene potentially linked with parental lifespan could not be identified in all genetic regions. This is the case for the *CELSR-PSRC1-SORT1, FURIN-FES, TXNL4B-HP-HPR* and *LAMA5-AL121832.2-CABLES2-CHRNA4* regions, within which the lead parental lifespan variant was linked with the expression of two or more genes. Multivariable MR, a MR technique used to identify the causal exposure accounting for potential confounders, did not identify the causal eGene from these loci (data not shown). We also observed that in the case of FURIN and HP, gene expression in one tissue was positively linked with parental lifespan while negatively linked with parental lifespan in another tissue. A Sankey diagram presents tissues underlying the genetic signals of colocalized eGenes (Fig. 2b). In order to gain insight into potential tissue specificity of the parental lifespan associated eGenes, we obtained the tissue-specific gene expression metric (Tau) as described by Kryuchkova-Mostacci and Robinson-Rechavi^7^. Genes with evidence of tissue-specific expression have a Tau value closer to 1 while ubiquitous genes have a Tau value closer to 0. This analysis revealed that several of the eGenes had tissue-specific expression (Tau ≥0.80), including the *HP* gene, which appears to be liver-specific, in accordance to our initial TWAS finding (Fig. 2c and Supplementary Table 2). Other tissue-specific eGenes include *KCNK3, NRG1, NEK10, CHRNA3/5* and *CHRNA4*. Visual inspection of single-cell sequencing data from human liver samples made available by Aizarani et al.^8^ further revealed a high expression of the *HP* gene within hepatocytes as opposed to other liver cell types (data not shown). We used LocusCompare^9^ to depict colocalization events within our framework and present as an example the colocalization of the top SNP associated with liver *HP* expression and parental lifespan (Fig. 2d).

**Figure 2.**
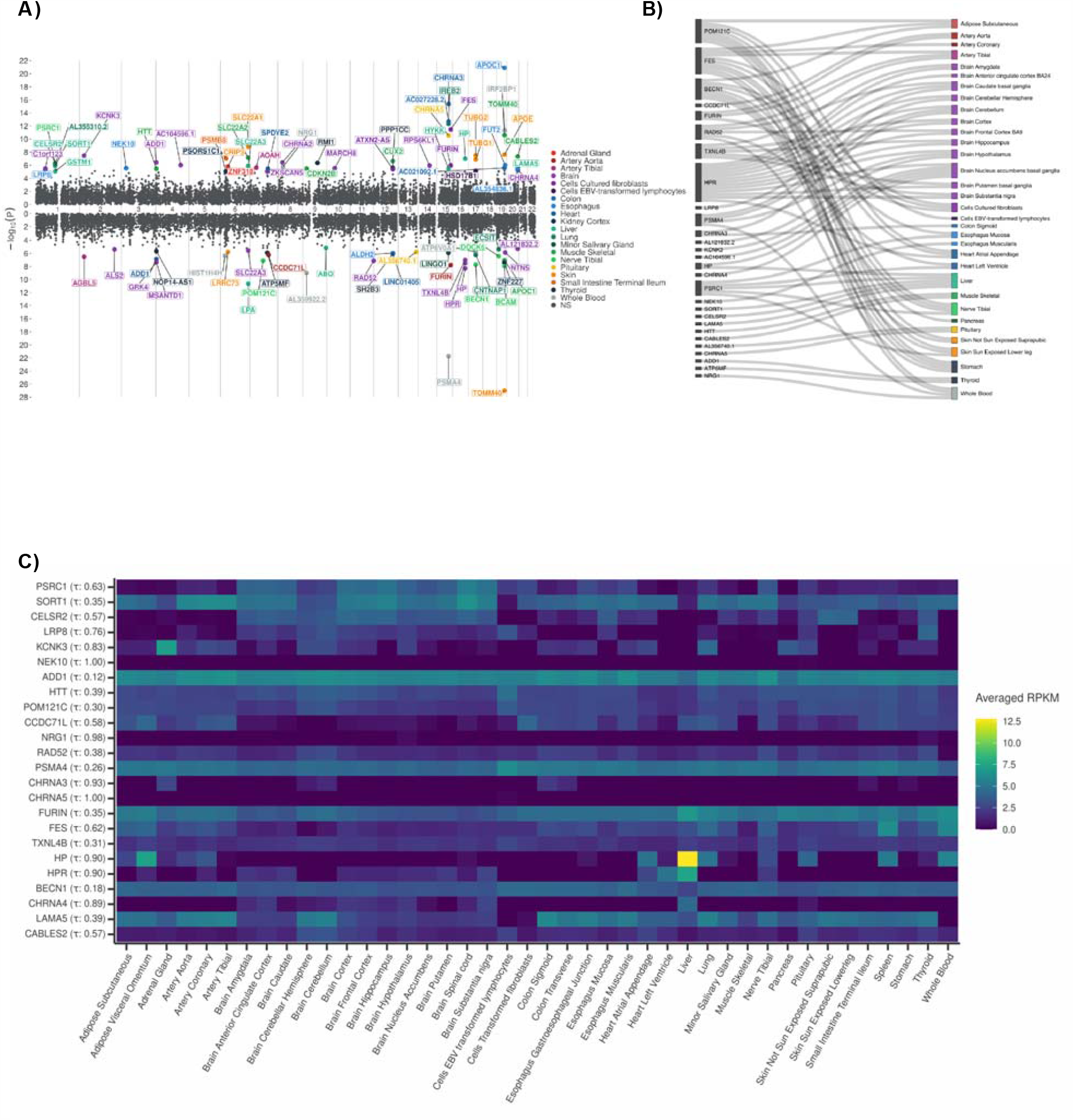
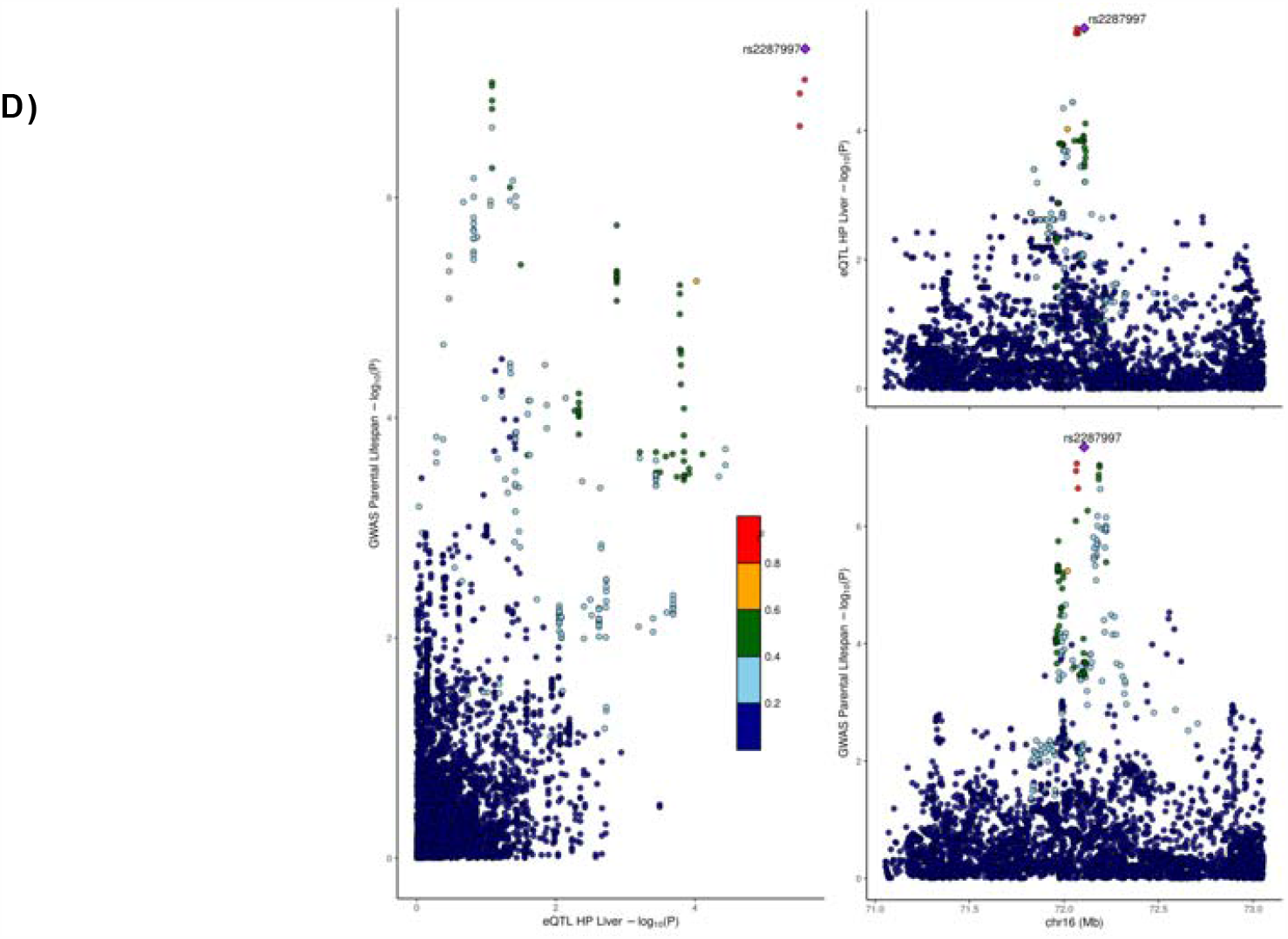
A multi-tissue transcriptome-wide association study of parental lifespan. A) Miami plot depicting the results of transcriptome-wide association studies of parental lifespan in multiple tissues before filtering out eGenes without evidence of genetic colocalization. Each dot represents the effect of an eGene on parental lifespan and the top tissue underlying the signal is shown. eGenes negatively associated with parental lifespan are above the baseline and eGenes positively associated with parental lifespan are below the baseline. Some tissues (for instance those in the brain) were pooled in the legend to facilitate visualization of the tissues responsible for the eGene-parental lifespan associations. B) Sankey diagram depicting tissues that are responsible for the eGene-trait associations. This analysis is based on 43 non-sex-specific tissues from GTEx. C) Heat map showing the tissue-specificity of the eGenes identified in this analysis (after filtering out eGenes without evidence of genetic colocalization). Tau value is shown in parentheses after the gene name. RPKM indicates reads per kilobase per million mapped reads. D) LocusCompare plot depicting colocalization of the top SNP associated with liver HP expression and parental lifespan.

**Table 1.**
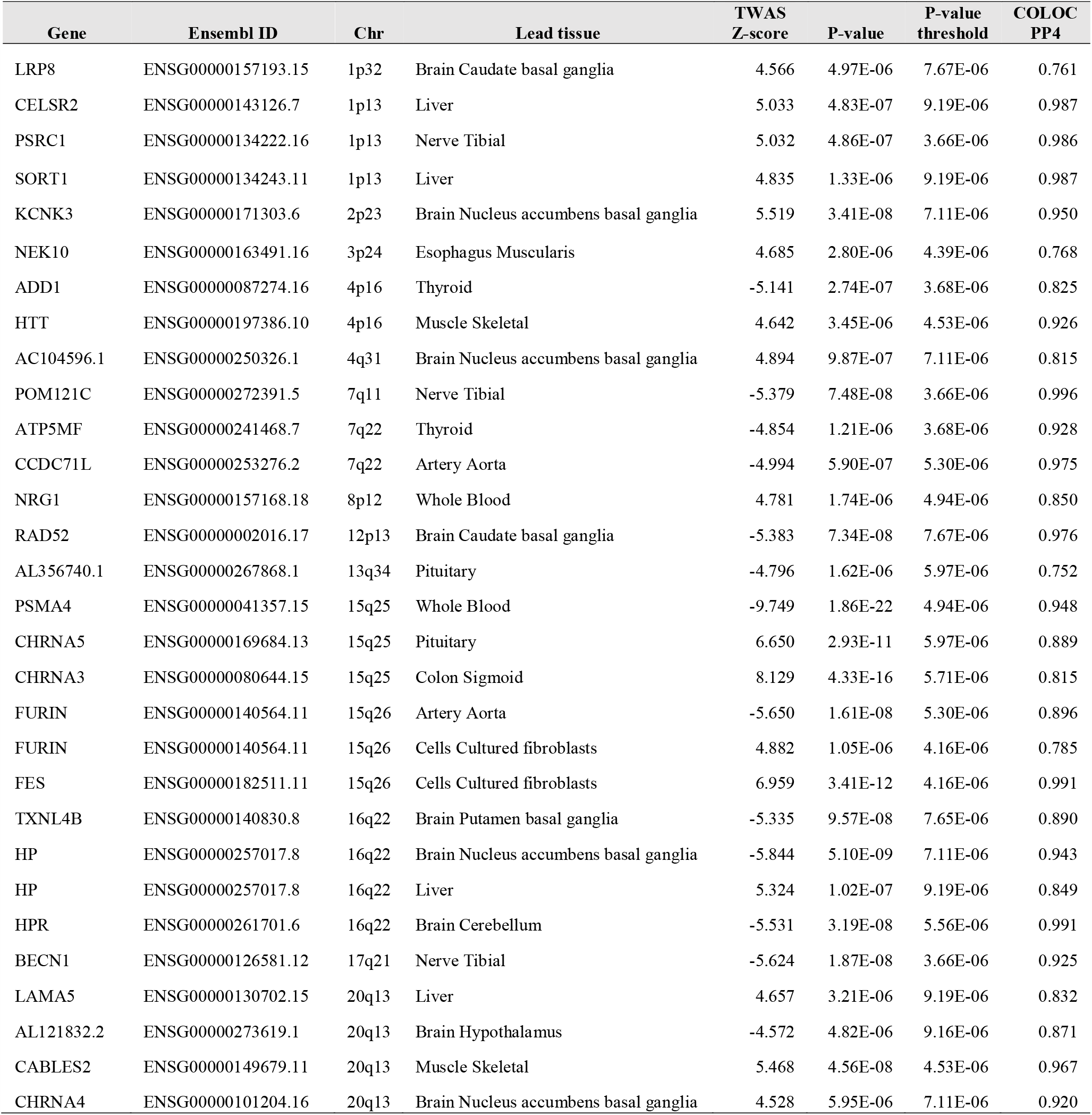
Significant eGene-parental lifespan associations from a transcriptome-wide association study of parental lifespan after filtering out eGenes without evidence of genetic colocalization.

### Identification of eProteins associated with parental lifespan

Proteins are the target of most medicines. In order to identify circulating factors causally linked with parental lifespan that could simultaneously represent therapeutic targets for human lifespan and healthspan, we performed a systematic screen of the human plasma proteome of the INTERVAL cohort^10^ using MR. We first obtained robust instruments for protein levels by identifying proteins with ≥4 cis-acting independent variants (r^2^<0.1) strongly associated with protein levels (p-value <5×10^−8^). A total of 279 circulating proteins were available for MR analyses using these criteria. We then used inverse variance weighted (IVW)-MR to determine the association between these circulating proteins and parental lifespan. After correction for multiple testing, nine proteins emerged as causal mediators of parental lifespan in the proteome-wide MR analysis, including haptoglobin (*HP* gene on chromosome 16q22), which has also been identified by GWAS and TWAS of liver-parental lifespan associations (Fig. 3A). Other proteins identified as causal mediators of parental lifespan include: asporin (*ASPN* gene on chromosome 9q22), agouti-signaling protein (*ASIP* gene on chromosome 20q11), the soluble receptor of insulin-like growth factor 2 (*IGF2R* gene on chromosome 6q25), plexin-B2 (*PLXNB2* gene on chromosome 22q13), interleukin-6 receptor subunit alpha (*IL6R* gene on chromosome 1q21), soluble intercellular adhesion molecule-5 (*ICAM5* gene on chromosome 19p13), ectonucleotidase phosphodiesterase 7 (*ENPP7* gene on chromosome 7q25) and Sushi, von Willebrand factor type A, EGF and pentraxin domain-containing protein 1 (*SVEP1* gene on chromosome 9q31). In order to determine whether these associations were confounded by pleiotropy and robust to outliers, we used additional outlier robust (MR-PRESSO) and modeling MR methods (MR-Egger and contamination mixture). The association of these proteins with parental lifespan using these methods are presented in Supplementary Table 3. Altogether, results of this analysis suggest that the causal association of these proteins with parental lifespan robust to outliers. The MR method however detected evidence of horizontal pleiotropy for one protein, IL6-sRA. Altogether, this analysis identified five circulating “protective” proteins that may be positively associated with parental lifespan and four circulating proteins that may be negatively associated with lifespan.

**Figure 3.**
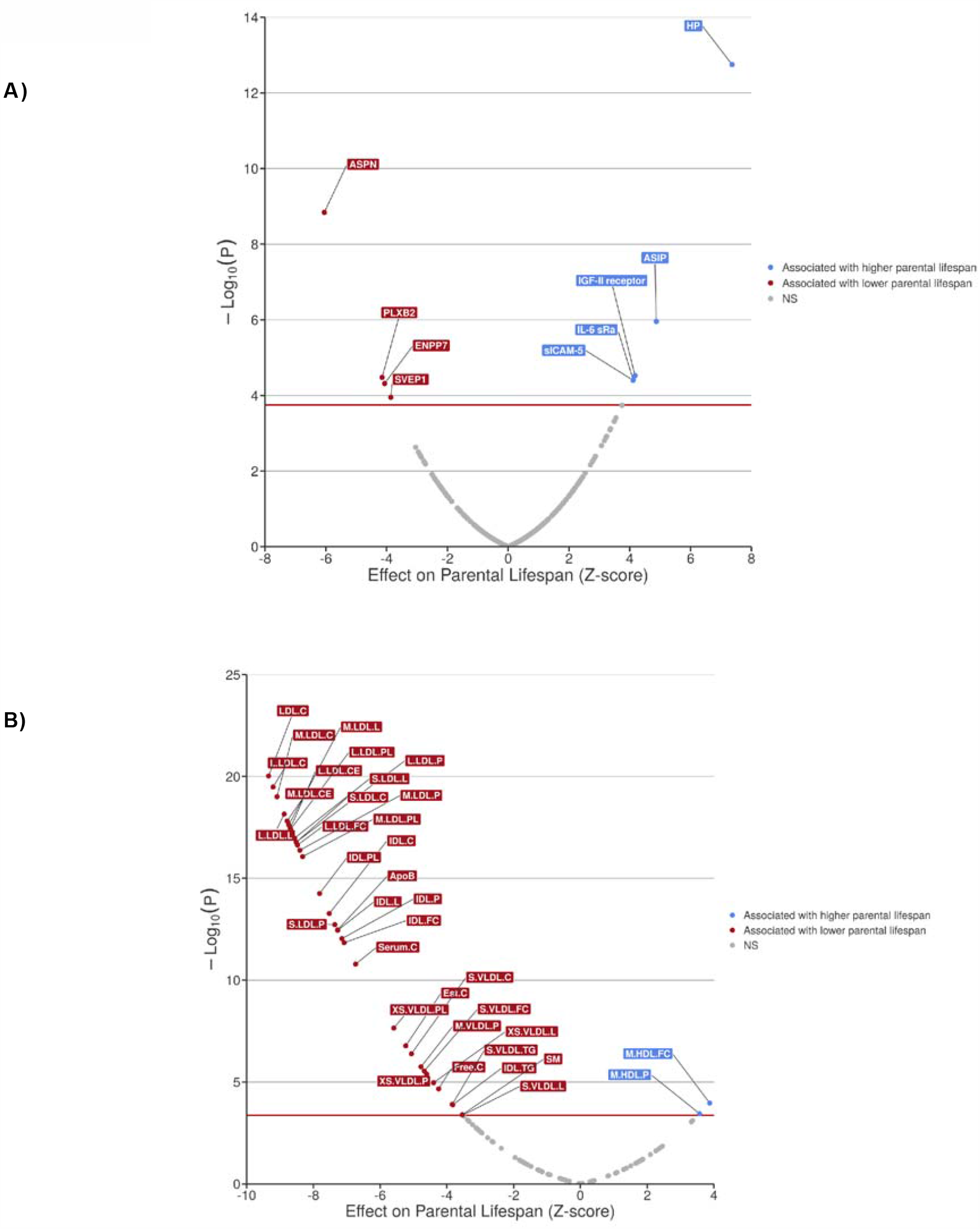
Proteome- and metabolome-wide Mendelian randomization analysis of parental lifespan. Volcano plots representing the association between plasma eProteins from the study of Sun et al. (A) and eMetabolites from Kettunen et al. (B) and parental lifespan using inverse-variance weighted Mendelian randomization. eProteins and eMetabolites in blue represent those positively associated with parental lifespan and those in red are negatively associated with parental lifespan.

### Identification of eMetabolites associated with parental lifespan

We applied a similar MR framework as above to identify eMetabolites including lipoprotein metabolomics parameters that could be causally associated with parental lifespan using a GWAS-metabolomics dataset, previously described by Kettunen et al.^11^ The association between 123 metabolites from this datasets and parental lifespan was investigated using IVW-MR (Fig. 3B). After correction for multiple testing, eMetabolites associated with parental lifespan included several parameters linked to very-low-density lipoprotein (VLDL) and low-density lipoprotein (LDL) physico-chemical properties (such as apolipoprotein B, cholesterol content and total VLDL/LDL particle number), sphingomyelin and the number of high-density lipoprotein particle of medium size (M:HDL-P). The same MR methods that enabled us to identify causal proteins associated with parental lifespan were used. Results of this analysis suggest that the causal association of these metabolites with parental lifespan are unlikely to be pleiotropic and robust to outliers (Supplementary Table 4). Altogether, results of this analysis suggest that circulating metabolites such as amino acids or glycolysis precursors may not be causally associated with parental lifespan while elevated levels apolipoprotein-B containing lipoproteins are strongly associated with shorter parental lifespan.

### Association of parental lifespan genetic instruments with age-related disease traits

We used summary statistics from 40 of the largest publicly available GWAS with at least 1000 cases, including 29 new meta-analyses that we performed using the UK Biobank and FinnGen studies (Supplementary Table 5)^12-20^ to determine the association between parental lifespan-associated eGenes-tissue pairs (Fig. 4A), eProteins (Fig. 4B), eMetabolites (Fig. 4C) and age-related chronic disease. Parental lifespan genome-wide significant variants that did not have TWAS associations (*MAGI3, CDKN2B-AS1, LPA, LDLR* and *APOE*), with the exception of *ATXN2*/*BRAP*, which we considered an eGene (*SH2B3*) as this gene passed the TWAS significance threshold with posterior probability of genetic colocalization of 0.63 were also included in this phase (Fig. 4D). Also, because eMetabolites causally associated with parental lifespan included many highly correlated parameters, we considered the top associated trait, LDL cholesterol level, as well as two other traits, sphingomyelin and HDL free cholesterol content. Altogether, we tested the association between 48 instruments (31 eGenes, 9 eProteins, 3 eMetabolites and 5 SNPs) and 40 outcomes. This analysis identified several trait-disease associations that were mostly concordant with their effect on parental lifespan (most traits associated with higher risk of disease were also associated with shorter parental lifespan). Detailed results of all statistically significant eGene/eProteins/eMetabolite/SNP-age-related disease trait associations tested in this analysis with multiple MR methods are presented in Supplementary Table 6, 7, 8 and 9, respectively. This analysis shows that although not all parental lifespan genetic regions were linked with age-related diseases, regions that were associated with age-associated diseases appeared to be mostly associated with diseases of the circulatory system.

**Figure 4.**
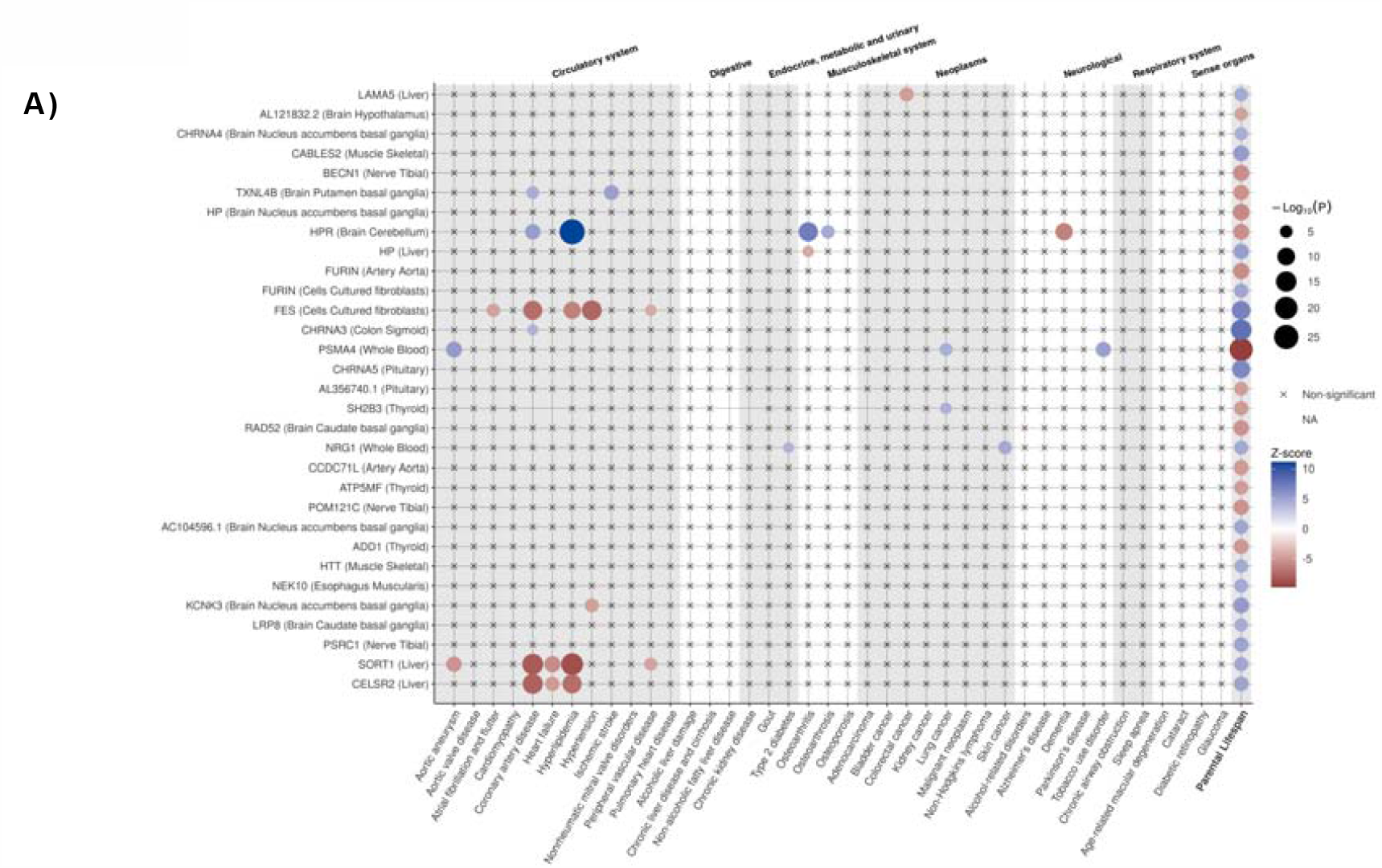
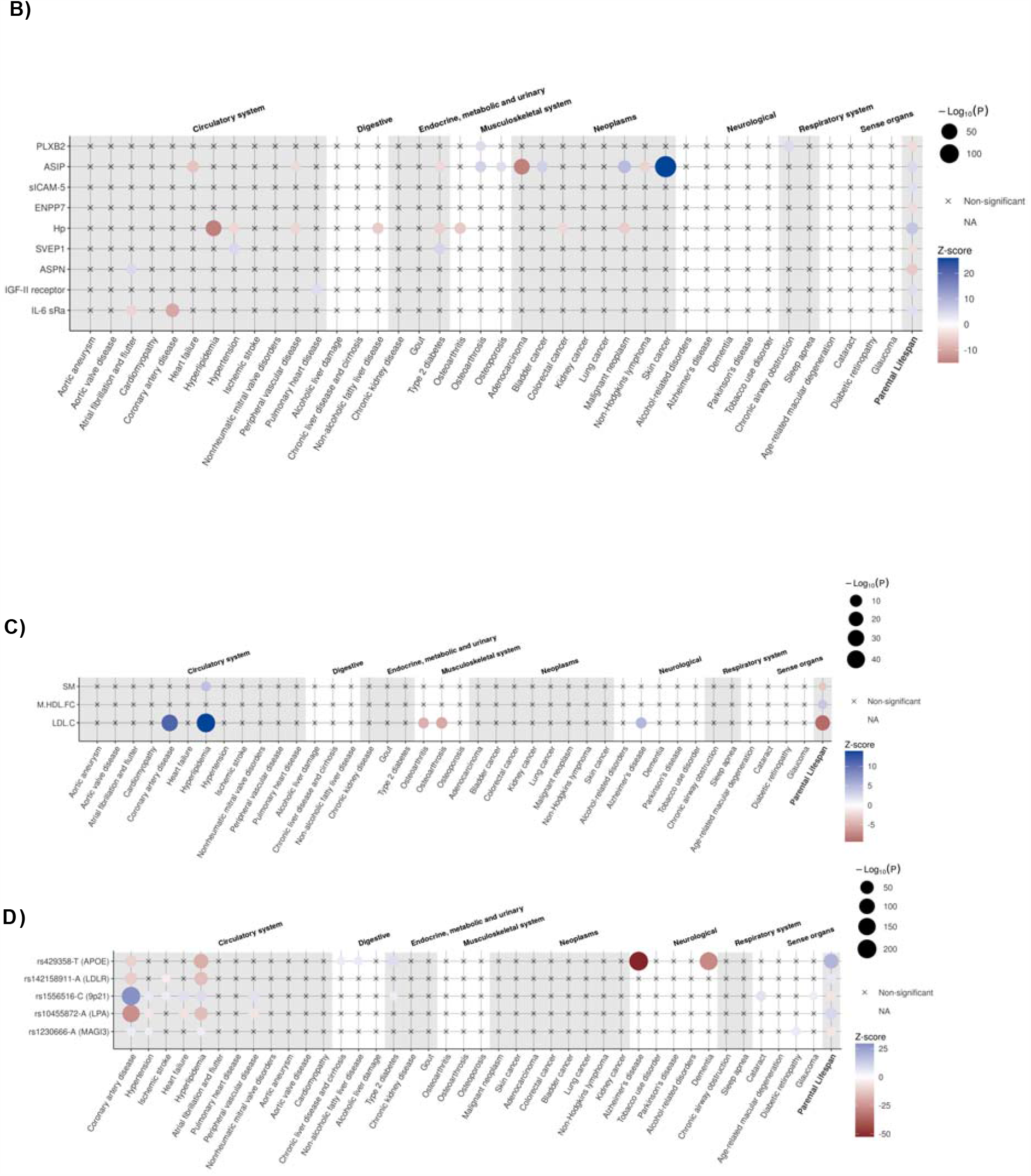
Mendelian randomization analysis of parental lifespan genetic determinants with 40 age-related chronic diseases. Balloon plots representing the association of A) eGenes, B) eProteins, C) eMetabolites from Kettunen et al., and D) parental lifespan variants not representing eGenes or eProteins with 40 age-related diseases or traits using inverse-variance weighted Mendelian randomization. A correction for multiple testing was applied.

### Investigation of shared genetic etiology at the HP locus

Given that the only parental lifespan signal that was supported by GWAS, TWAS and proteome-wide MR was at *HP*, the gene encoding haptoglobin (Hp) and that variation at the HP locus was linked with hyperlipidemia, we investigated shared genetic etiology across these traits. For this purpose, we used a Bayesian algorithm called Hypothesis Prioritization in multi-trait Colocalization (HyPrCOLOC)^21^. In order to reduce the risk associated with spurious association from overlapping dataset, we used GWAS summary statistics from the Global Lipids Genetics Consortium for LDL-C. Results presented in Fig. 5a suggest shared genetic etiology between liver *HP* expression, circulating LDL levels, and parental lifespan. Upon further investigation, we found that liver *HP* expression, circulating LDL levels, parental lifespan but not circulating Hp levels likely shared genetic etiology at this locus with a posterior probability of colocalization of 0.96 Fig. 5b.

**Figure 5.**
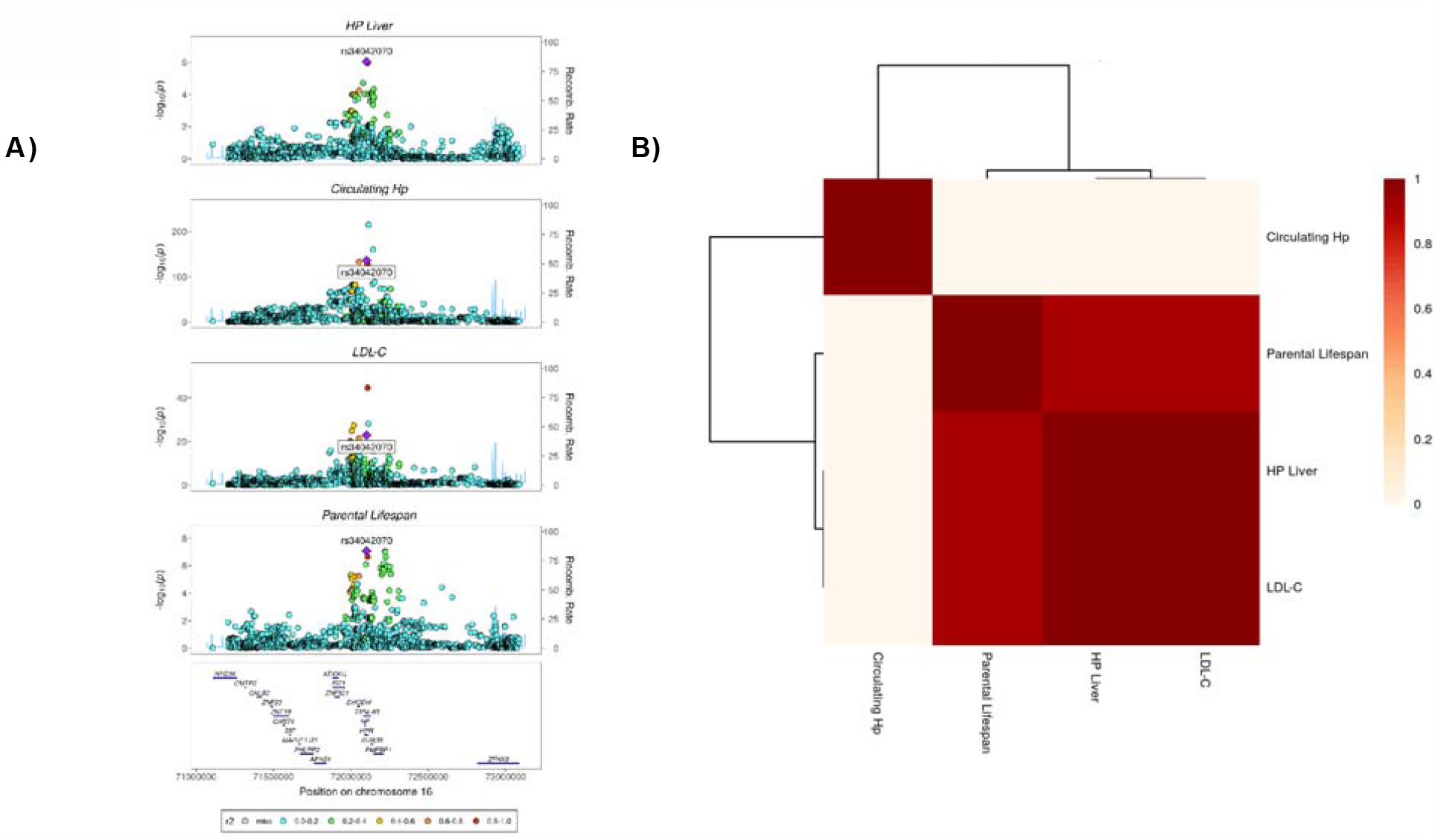
Statistical colocalization at the HP locus. A) Regional association plot highlighting the lead SNP associated with liver HP expression (rs34042070) and circulating haptoglobin levels, low-density lipoprotein cholesterol levels and parental lifespan with color key indicating r^2^ with the lead SNP. B) Heatmap depicting the posterior probabilities of colocalization between each trait pair.

### Identification of drug repositioning opportunities for human lifespan/healthspan extension

As many of the eGenes, eProteins and eMetabolites were associated with age-related chronic disease, we aimed at identifying in a more comprehensive manner which disease traits might be influenced by these exposures. We performed a systematic analysis of potential diseases associated with eGenes and genes encoding eProteins in the Open Targets Platform^22^. eMetabolites were not considered in this analysis as their variance is determined by more than one gene each contributing to eMetabolite levels to a different extent. Supplementary Table 10 presents the diseases and therapeutic areas relevant to the identified eGenes and eProteins with p-values <5×10^−8^ for association score with many therapeutic areas concordant with the effect of eGenes and eProteins on chronic disease (Table 1 and Fig. 3). We further searched the Open Targets Platform to identify drug candidates targeting eGenes and eProteins to identify possibly drug reposition opportunities for lifespan/healthspan extension. Supplementary Table 11 presents 23 drug candidates that were identified by Open Targets Platform. These drugs target four parental lifespan eGenes (*CHRNA4, KCNK3, CHRNA3/5*, and *HTT*) and one parental lifespan eProtein (*IL6R*). A second strategy was also used to intersect eGenes and eProteins with the DrugBank and the Drug Gene Interaction database^23^. This analysis identified new targets potentially interacting with parental lifespan eGenes and eProteins (Supplementary Table 12 and 13). Finally, we used the network processing data tool GeneMANIA to identify new genes that may show coexpression or physical interaction with our targets^24^. The inner circle of Fig. 6a presents includes the eGenes and genes encoding for eProteins that we have entered in the engine while the outer circle presents new targets that show coexpression or physical interaction with our targets. Consistent with the analysis of Timmers et al.^4^, this analysis also enabled the identification of the top biological pathways (all with FDR p-value<1×10^−4^), which are all related to lipoprotein metabolism.

**Figure 6.**
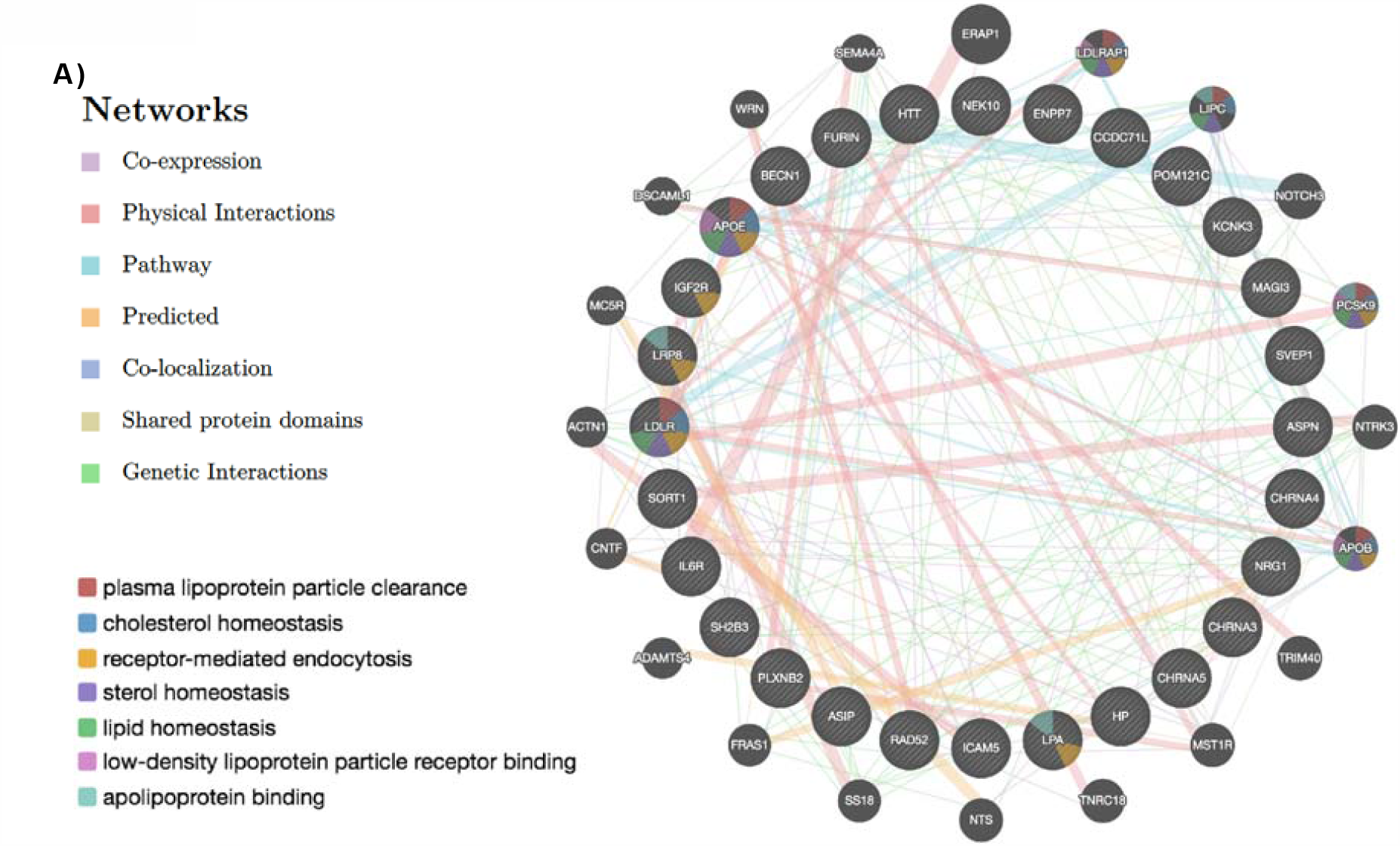
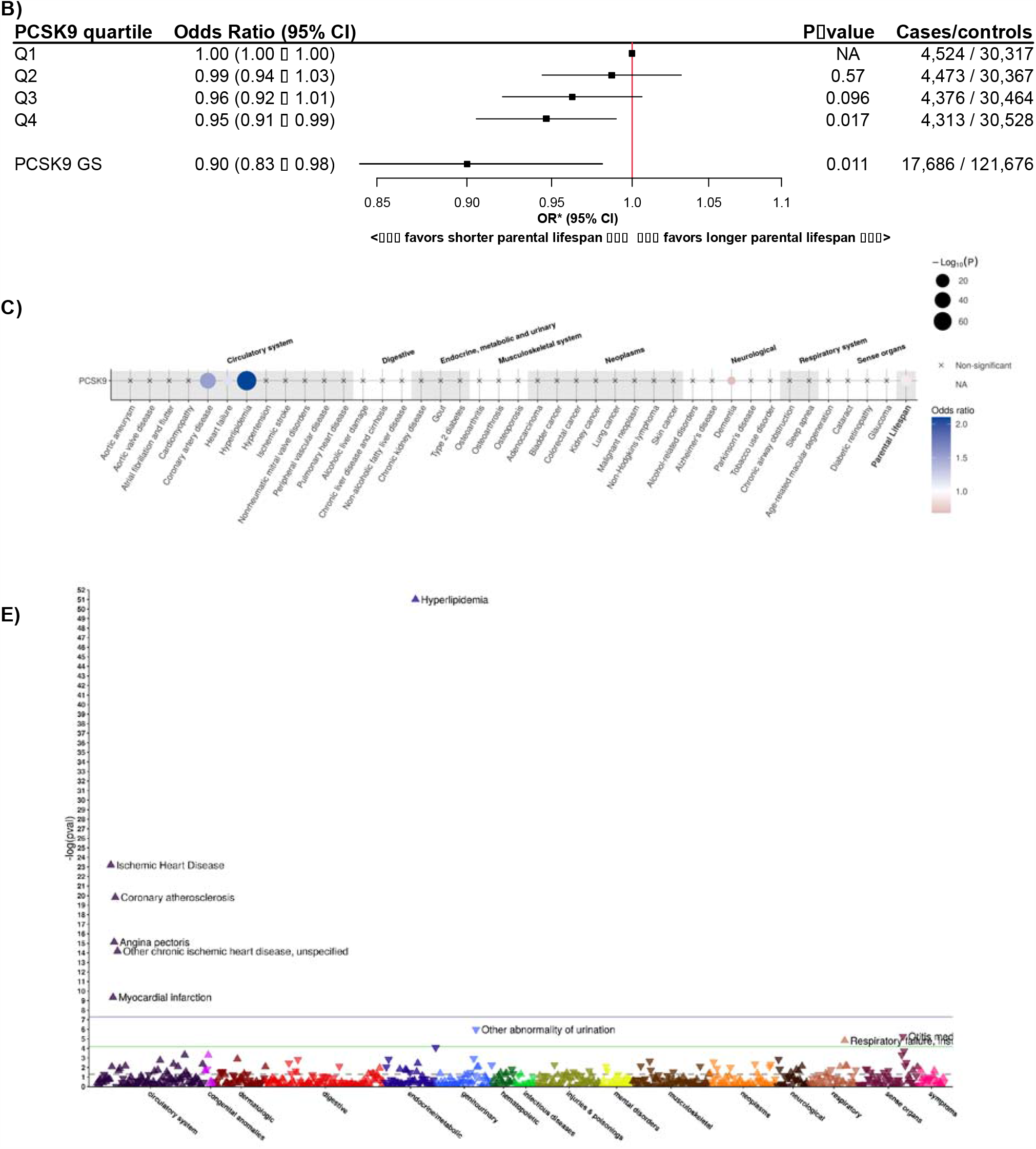
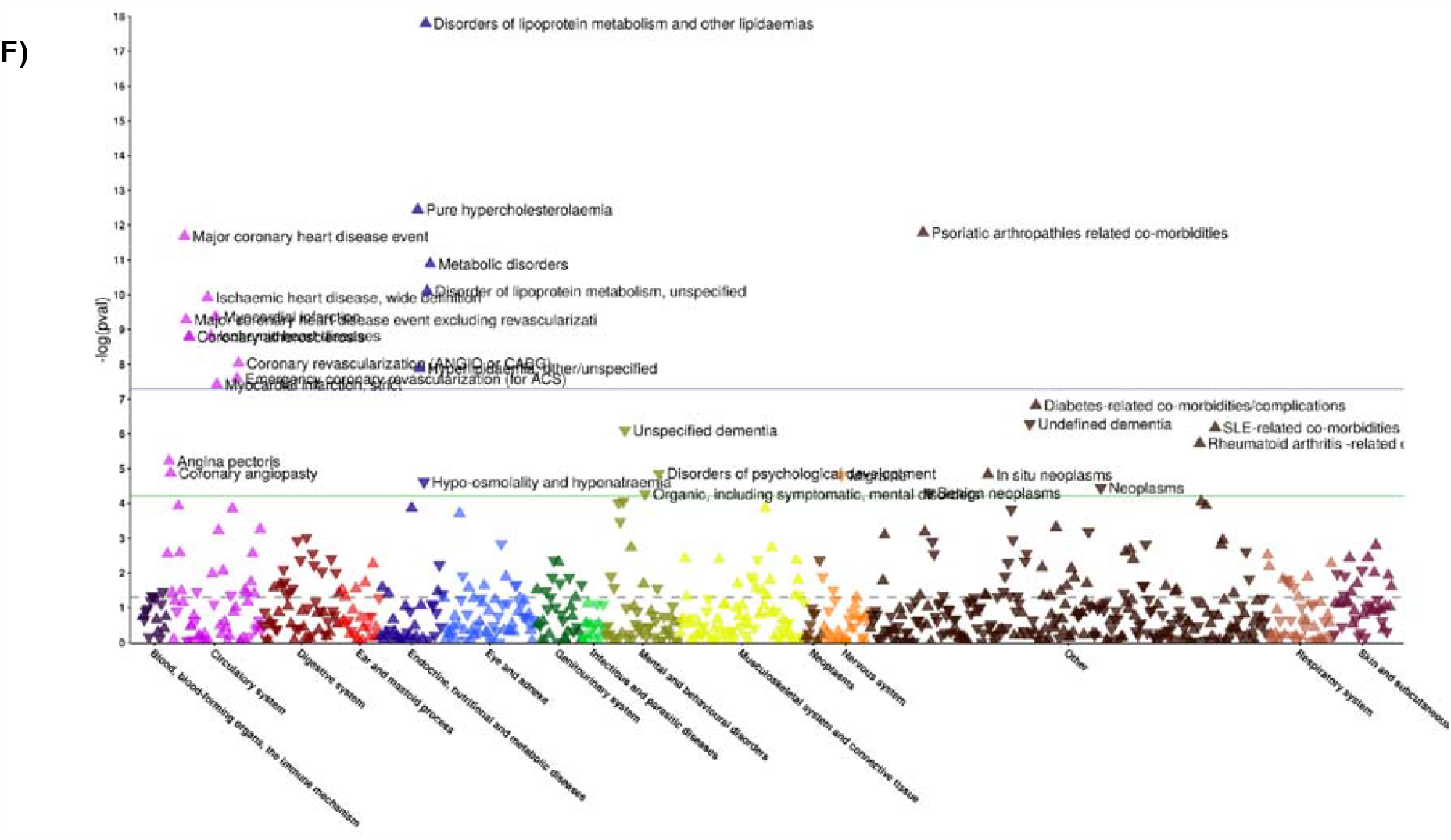
Investigation of proprotein convertase subtilisin/kexin type 9 (PCSK9) inhibition as a potential human healthspan extension therapy. A) Interaction network of parental lifespan eGenes and eProteins with other genes and top pathways of parental lifespan eGenes and eProteins (false discovery rate <5×10^−5^) from the GeneMANIA prediction server. B) Odds ratio (OR) and 95% confidence interval (CI) for high parental lifespan in participants of the UK Biobank separated into quartiles of the PCSK9 genetic score (GS) of 11 independent variants associated with low-density lipoprotein cholesterol levels. Analyses were adjusted for age, sex and 10 first ancestry-based principal components. The adjusted odds ratio and 95% CI for high parental lifespan, associated with a 1 mmol/L increase in low-density lipoprotein cholesterol associated with these variants is also shown. C) Balloon plots representing the association between variation at the PCSK9 locus with 40 age-related diseases or traits and parental lifespan. A total of 11 independent SNPs at the PCSK9 locus were weighted for the impact on low-density lipoprotein cholesterol levels and Mendelian randomization was performed using inverse-variance weighted Mendelian randomization. Associations are reported after correction for multiple testing (p-value <0.05/41 [<1.22×10^−3^]). D) Phenome-wide inverse-variance weighted Mendelian randomization study depicting the association between variants at the PCSK9 locus weighted for their impact on low-density lipoprotein cholesterol at 791 binary disease-related traits in the UK Biobank and 809 binary disease-related traits in FinnGen. E) Associations are reported after correction for multiple testing in the UK Biobank (p-value <0.05/791 [<6.32×10^−5^]) and FinnGen (p-value <0.05/809 [<6.18×10^−5^]). Arrows pointing up represent higher disease presence and arrows pointing down represent lower disease presence. The dotted line represents the nominal p-value of 0.05, the green line represents the p-value after correction for multiple testing and the blue line represents the genome-wide significance level.

### Genetic investigation into PCSK9 as a therapeutic target for human healthspan extension

Proprotein Subtilisin/Kexin Type 9 (PCSK9) emerged as a potential drug target for several reasons. First, PCSK9 is involved in the top 6 pathways suggested by GeneMANIA. Second, GeneMANIA suggested co-expression with *LDLR*, a parental lifespan locus. Experimental evidence supports this finding since both PCSK9 and the LDLR are under sterol regulatory element-binding protein 2 control^25^. Third, PCSK9 interacts with apolipoprotein B, and an important proportion of circulating PCSK9 is carried by apolipoprotein B-containing lipoproteins^26^ including lipoprotein(a), another parental lifespan locus, *LPA*^27^. Fourth, PCKS9 also has shared proteins domains with another parental lifespan eGene, *FURIN* (a finding that is also supported by experimental evidence since furin has the ability to cleave PCSK9^28^). Fifth, DrugBank and the Open Targets Platform suggested PCSK9 neutralizing antibodies (alirocumab and evolocumab) as potential drug repurposing opportunities. Sixth, PCSK9 inhibitors have important LDL cholesterol reduction properties (up to 60%). As our investigation reported a strong and causal effect of exposure to low LDL cholesterol levels and parental lifespan (Fig. 3b), we investigated the association between genetic variation at the *PCSK9* locus and parental lifespan using individual participant level data in the UK Biobank. We identified 11 independent SNPs within the *PCSK9* region strongly and independently associated with LDL cholesterol in the Global Lipids Genetic Consortium (r^2^<0.1 and p-value<5×10^−8^). We evaluated the impact of carrier status of *PCSK9* SNPs linked with high LDL and the impact of a weighted genetic score (GS) scaled to model a 1 mmol/L increase in LDL-C levels and found a significant association with having lower odds of high parental lifespan (Fig. 6b). Investigating the association between the GS and the previously reported 40 age-related disease traits, we found that variation in *PCSK9* associated with higher LDL cholesterol levels was associated with shorter parental lifespan, higher risk of hyperlipidemia, coronary artery disease and heart failure and potentially associated with a lower risk of dementia after correction for multiple testing (Fig 6c). We next performed a phenome-wide MR analysis across the disease-related phenome to gain more insight on the potential benefits/risk ratio of potential human lifespan extending drugs such as PCSK9 inhibitors. The association between 11 SNPs at the *PCSK9* locus (weighted for their impact on plasma LDL cholesterol levels) and 791 binary disease-related traits in 408,961 participants of the UK Biobank was investigated. As expected, PCSK9 “genetic inhibition” may increase healthspan through its association with lifelong exposure to low LDL cholesterol levels and associated reduction in the risk of cardiovascular diseases (Fig. 6d and Supplementary Table 14). Because there was marked participant overlap in the cohorts evaluating both the study outcomes (parental lifespan UK Biobank and LifeGen consortium) and disease-traits (UK Biobank only, we investigated association between the same genetic instrument and 809 binary disease-related traits in 96,499 participants of the FinnGen cohorts (Fig. 6e and Supplementary Table 15). This analysis confirmed the high impact of *PCSK9* variants on cardiovascular diseases. This analysis also revealed an association between higher circulating LDL cholesterol levels associated with *PCSK9* variants and lower dementia in FinnGen, but not in the UK Biobank.

### Phenome-wide Mendelian randomization studies across the disease-related phenome

The eGenes, eProteins and eMetabolites that showed robust association with our longevity trait could represent potential therapeutic targets for increased lifespan/healthspan. In order to gain knowledge on the function of these genes and proteins, and to predict potential benefits and/or adverse effects of therapeutic agents that could be developed to mimic the effect of eGenes, eProteins eMetabolites and other genetic regions of human health, we performed a phenome-wide MR across the disease-related phenome. We used IVW-MR with genetic instruments for eGenes, eProteins, eMetabolites and other genetic regions to perform MR-pheWAS for 791 disease-specific binary traits in the UK Biobank and for 809 disease-specific binary traits in the FinnGen cohorts. Details on all diseases that passed correction for multiple testing using MR-pheWAS analyses in the UK Biobank are presented in Supplementary Tables 16, 17, 18 and 19 for parental lifespan eGenes, eProteins, eMetabolites and other parental lifespan SNPs, respectively. This analysis yielded 636 gene-disease trait associations. MR-pheWAS analyses in the FinnGen cohorts are presented in Supplementary Tables 20, 21, 22 and 23 for parental lifespan eGenes, eProteins, eMetabolites and other parental lifespan SNPs, respectively. A total of 493 gene-disease trait associations were reported in FinnGen.

### Candidate drugs for human healthspan extension

Finally, we combined results from our genetic instruments (eGenes, eProteins, eMetabolites and other parental lifespan variants), the candidate drugs identified by our search in the Open Target Platform, DrugBank, the Gene-Drug interaction database, a literature search of other drugs under development and our targeted (40 age-associated diseases) and untargeted (phenome-wide MR in UK Biobank and FinnGen) approaches to propose a list of potential drug targets for human healthspan extension. Results from this analysis presented in Table 2 and suggest many novel and known pathways that may be investigated in randomized clinical trials to enhance human healthspan.

**Table 2.**
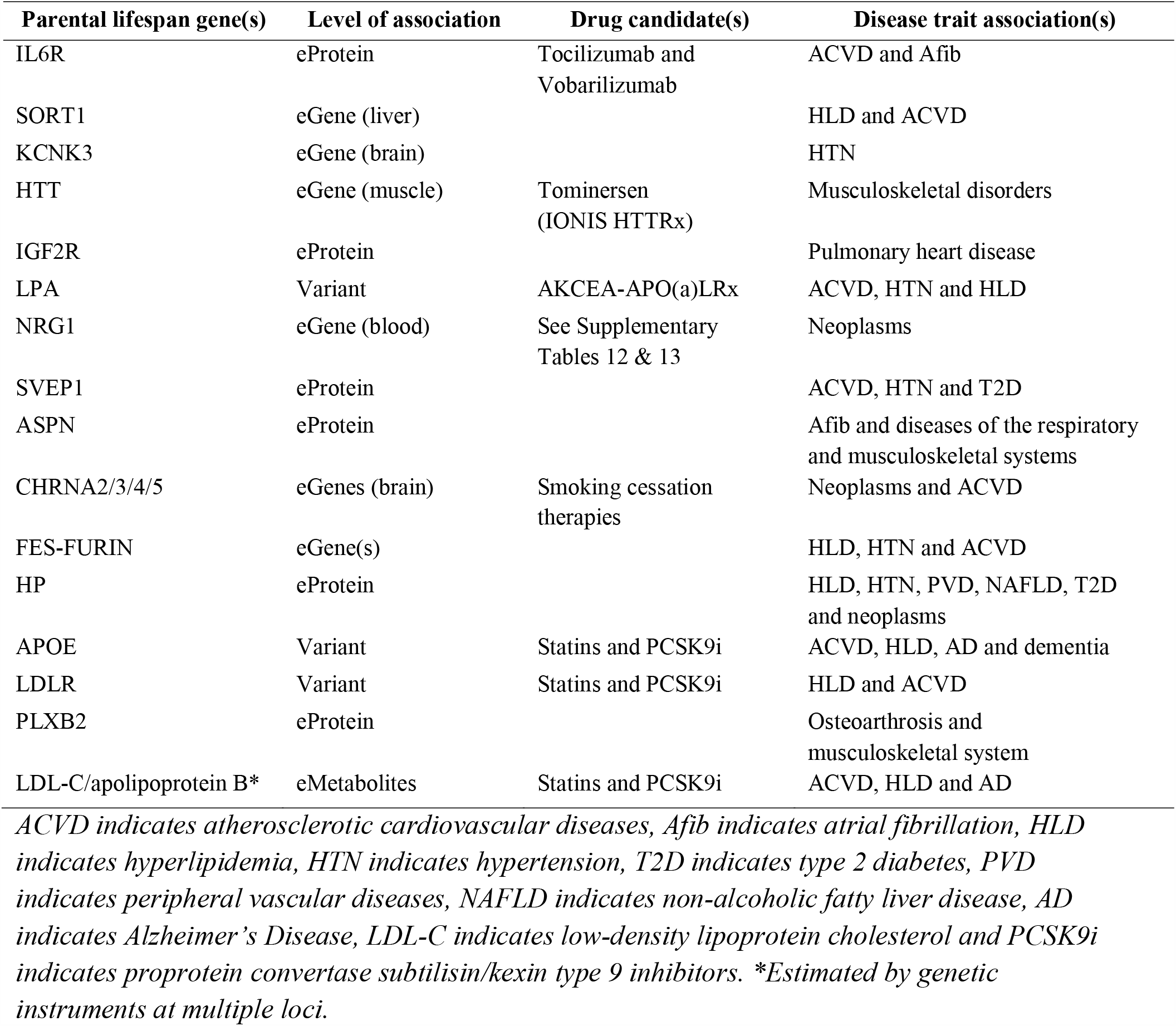
Candidate drug targets for healthspan extension identified by trans-omic Mendelian randomization study of parental lifespan.

## DISCUSSION

Our translational MR approach that combined TWAS with genetic colocalization, proteome-wide, metabolome-wide and phenome-wide MR sought to identify novel biological determinants of parental lifespan and their relevance in age-associated human diseases as well as new therapeutic targets to promote healthspan. The newly identified genomic loci colocalizing with parental lifespan include *LRP8*, the gene encoding the low-density lipoprotein receptor-related protein 8 (also known as ApoER2), *NEK10*, the gene encoding NIMA related kinase 10, *CCDC71L* the gene encoding Coiled-coil domain-containing protein 71L), *NRG1*, the gene encoding neuregulin-1, *RAD52*, the gene encoding DNA repair protein RAD52 homolog and *BECN1*, the gene encoding beclin-1, the latter being also reported in our previous MR study of parental lifespan using blood gene expression as instrumental variables)^29^. Our approach strengthens the case for previously identified regions by GWAS by the identification of a tissue-specific colocalized TWAS signal for the *KCNK3, ADD1*/*GRK4*/*HTT, HP*/*HPR*/*TXNL4B, LAMA5/ AL121832.2/CABLES2/CHRNA4* loci. We provide evidence using genetic colocalization that genetic regions previously identified by TWAS (*CELSR2*/*PSRC1*/*SORT1, POM121C, CHRNA3/5* and *FES/FURIN*) are likely causally associated with parental lifespan through shared genetic etiology. Results of our analysis also suggest tissue-specific regulation of parental lifespan eGenes consistent with a biological effect. Some examples include brain eGenes such as *LRP8* and smoking-associated eGenes (*CHRNA2/3/4/5*), liver eGenes such as *HP* and *SORT1*, and a skeletal muscle eGene (*HTT*). Finally, we provide evidence that nine circulating proteins (haptoglobin [*HP*], asporin [*ASPN]*, agouti-signaling protein [*ASIP]*, the soluble receptor of insulin-like growth factor 2 [*IGF2R]*, plexin-B2 [*PLXNB2]*, interleukin-6 receptor subunit alpha [*IL6R*], soluble intercellular adhesion molecule-5 [*ICAM5*], ectonucleotidase phosphodiesterase 7 [*ENPP7*] and Sushi, von Willebrand factor type A, EGF and pentraxin domain-containing protein 1 [*SVEP1*] may be causally associated with parental lifespan. Many of these blood factors could represent therapeutic targets to reduce the risk of age-associated chronic diseases and longevity. Our search for parental lifespan eMetabolites revealed that circulating metabolites such as amino acids or energy metabolism associated molecules may not be causally associated with parental lifespan. However, plasma levels apolipoprotein-B containing lipoproteins (VLDL and LDL) were strongly associated with shorter parental lifespan. This MR analysis provides genetic confirmation of the results several investigations into the long-term health benefits of lipid-lowering therapies^30,31^.

In support of the causal association of apolipoprotein-B-containing lipoprotein and parental lifespan, we found through a gene interaction network that the four genes responsible for the overwhelming majority of the cases for the most prevalent human genetic disease^32^, familial hypercholesterolemia, could be linked with genes regulating lifespan (*LDLR, PCSK9, APOB* and *LDLRAP1*). These results support the notion that lifelong exposure to low concentration of all apolipoprotein-B-containing lipoprotein particles might be an important determinant of atherosclerotic coronary artery diseases and human longevity. In this work, we also provide evidence that genetic variation in *PCSK9* mimicking the effect of PCSK9 inhibitors is linked to lifelong exposure to low LDL cholesterol levels and higher parental lifespan. These results suggest that PCSK9 inhibitors could be key to reduce LDL cholesterol and extending lifespan/healthspan if tested in adequately powered long-term randomized clinical trials. Unexpectedly, we found an association between reduced *PCSK9* function and higher risk of dementia (a finding observed in the FinnGen cohorts, but not in the UK Biobank). A previous large MR study found no association between variation in *PCSK9* and dementia^33^ and another found an association between variants linked with higher *PCSK9* function and Alzheimer’s Disease (AD) risk, and higher parental lifespan^34^. This finding might also be due to survival bias of individuals exposed to lifelong lower levels of LDL cholesterol. PCSK9 might also interact with *LRP8* (*APOER2*), a gene identified here as a potential regulator of parental lifespan for the first time to our knowledge.

A non-colocalized signal was also reported for a liver expressed gene: *LPA*, the gene that encodes apolipoprotein(a), which is one of the components of lipoprotein(a) (Lp[a]), another apolipoprotein-B-containing highly atherogenic lipoprotein particle^35^. We have previously reported in a hypothesis-driven MR study a strong relationship between plasma Lp(a) levels and parental lifespan, healthspan (defined as disease-free survival) and long-term mortality risk^36^. Long-term health outcomes trials of lipoprotein(a)-lowering therapies with antisense oligonucleotides are currently underway (NCT04023552).

In their landmark study investigating the genetics of parental lifespan in more than one million individuals, Timmers et al.^4^, developed and used a new approach called Bayesian prior-informed GWAS to identify new genetic variants associated with parental lifespan based on mortality risk factors. This approach led to the discovery of 7 loci associated with parental lifespan. Our trans-omic MR study identified 3 of these loci (without using prior information on mortality risk factors): *IGF2R* using proteome-wide MR as well as *POM121C* and the *CELSR2*/*PSRC1*/*SORT1* region using TWAS. Although the same genetic variant was responsible for the genetic colocalization of the three co-regulated genes at the 1p13 locus (*CELSR2*/*PSRC1*/*SORT1* region), *SORT1* might be responsible for the parental lifespan signal at the 1p13 locus. Indeed, a large body of evidence supports a functional impact of sortilin (encoded by the *SORT1* gene) in lipoprotein metabolism^37,38^. Altogether, these results support the usefulness of combining of various genetic investigation techniques relying on mortality risk factors and MR to identify new genetic and biological determinants of complex traits such as human longevity.

Results of this study further confirm the role of smoking behaviors as a strong determinant of lifespan and healthspan. We found that many smoking-associated loci such as cholinergic receptor nicotinic 2, 3/5 and 4 subunits may actually be brain eGenes. Timmers et al.^4^ did identify variation at *CHRNA 3/5* on chromosome 15 to be associated with parental lifespan at the genome-wide significance level. Here we report that *CHRNA4* on chromosome 20 and *CHRNA2* on chromosome 8 as new potential parental lifespan loci (although the latter did not show evidence of genetic colocalization). Unsurprisingly, our search for potential healthspan extension drug targets identified many smoking cessation therapies and the results of our phenome-wide MR studies confirmed the association of these eGenes with atherosclerotic cardiovascular diseases and neoplasms. These results confirm the importance of urgently adopting strict tobacco control policies throughout the world to improve global health, healthspan and longevity.

In this analysis, the only protein that showed significant association in genome-wide, transcriptome-wide and proteome-wide MR studies was haptoglobin (Hp). We show here that *HP* is a liver expressed gene and that liver expression of *HP*, as well as plasma Hp is linked with parental lifespan. A study showed that Hp levels are higher in the plasma of Japanese semisuper centenarians^39^. Hp is an acute phase protein and is one of the most abundant protein in human plasma. It binds free hemoglobin and facilitates its removal from the bloodstream. Hp also decreases oxidation of apolipoprotein E (encoded by the longevity gene *APOE*). A previous study showed that exon deletion in *HP* was associated with lower LDL cholesterol levels^40^. The authors of that study proposed a mechanism whereby Hp reduced the oxidation of apolipoprotein E and facilitated its removal from the blood stream, thereby reducing LDL cholesterol levels. Upon investigation of shared genetic etiology at the HP locus on chromosome 15, we found strong evidence of genetic colocalization between liver HP expression, LDL-C levels and parental lifespan, but not plasma Hp levels. We believe that this finding might be due to the fact that the assessment of plasma Hp levels may not capture the full extent of the various circulating Hp isoforms (1-1, 1-2 and 2-2). Interestingly, our gene network analysis revealed a potential interaction of HP and APOE. Interaction of Hp, APOE and beta-amyloid was also reported in human brain tissues, where this complex might influence central cholesterol metabolism^41^. This finding is also supported by our phenome-wide MR study in FinnGen reporting a positive association between liver expression of *HP* (but not blood levels of Hp) and AD. Functional genomics and experimental investigations will be needed to shed light on the mechanisms linking *HP* and longevity and to determine if *HP* might represent a potential therapeutic target to promote healthspan.

The majority of previous investigations into the biological determinants of aging have contributed to the identification of several biological mechanisms regulating longevity in model organisms. These mechanisms include, amongst others, the modulation of the insulin-like signalling pathway, target of rapamycin, sirtuins and NAD+, circadian clocks, mitochondria and oxidative stress, senescence, chronic inflammation, autophagy, proteostasis as well as genomic instability and telomere attrition^42^. Although the biological determinants of aging in humans are still incompletely understood, aging mechanisms identified in model organisms appear to contrast with the biological determinants of aging in humans. One exception, however, appears to be chronic inflammation, which might influence longevity in both model organisms and humans, as supported by our recent report of genetic variation in the interleukin (IL)-6 signalling pathway being associated with parental lifespan^43^ and the results of the CANTOS trial, which reported important mortality benefits in patients with elevated high-sensitivity C-reactive protein levels following administration of the IL-1β monoclonal antibody canakinumab^44^. Our study builds on this previous work showing that genes involved in both innate and immune response may be associated with longevity^29^. Here, our proteome-wide MR study identified variation at the *IL6R* and *ICAM5* loci to be associated with parental lifespan. Although some cardiovascular benefits could be observed by targeting the IL-6 or ICAM5 pathways, our phenome-wide MR results suggest that this might come at the price of increased risk of atopic disorders.

Other noteworthy findings of our study include a possible interaction between a newly identified locus, *RAD52* and *WRN*, the Werner syndrome ATP-dependent helicase. Coordination of these two proteins activities involves telomere metabolism through their action on replication fork rescue after DNA damage in cells of patients with Werner syndrome (a premature aging disorder). Another finding of interest is the identification of Sushi, von Willebrand factor type A, EGF and pentraxin domain-containing protein 1 (SVEP1) as a circulating protein causally associated with parental lifespan. Variation in this gene has been linked with hypertension and cardiac dysrhythmias^45^ and MI^46^, a finding replicated in our phenome-wide MR analysis in the UK Biobank. Interestingly, a previous analysis also reported a potential association in this gene with longevity in the Framingham Heart Study^47^.

In conclusion, our study identified new genetically regulated genes across 43 tissues, as well as genetically regulated circulating proteins and metabolites that potentially regulate human lifespan. Many of these genetic determinants of parental lifespan may represent potential therapeutic targets for human healthspan extension. Our study also underscores the importance of global population health measures such as adopting stricter tobacco control measures as well as the globalization of interventions targeting all apolipoprotein-B containing particles to prevent the onset of diseases of the cardiovascular system and possibly promote longevity.

## Data Availability

URLs for external datasets and statistical software are included in the manuscript.

## ACKNOWLEDGEMENTS

We would like to thank all study participants as well as all investigators of the studies that were used throughout the course of this investigation (LifeGen consortium, UK Biobank, FinnGen, INTERVAL, etc.). NP holds a doctoral research award from the *Fonds de recherche du Québec: Santé*. (FRQS). BJA and ST hold junior scholar awards from the FRQS. PM holds a FRQS Research Chair on the Pathobiology of Calcific Aortic Valve Disease. YB holds a Canada Research Chair in Genomics of Heart and Lung Diseases. The Genotype-Tissue Expression (GTEx) Project was supported by the Common Fund of the Office of the Director of the National Institutes of Health, and by NCI, NHGRI, NHLBI, NIDA, NIMH, and NINDS.

## METHODS

### Multi-tissue transcriptome-wide association study of parental lifespan

The main study outcome was parental survival, which was obtained from summary statistics of the study of Timmers et al.^4^ who performed a GWAS of survival in a sample of 1,012,240 parents from the UK Biobank (691,621 parental lifespans) and cohorts of the LifeGen consortium (320,619 parental lifespans from 26 additional population cohorts). This study identified parental lifespan was defined by the age of parent death or dead/alive status. Parents with an age of death <40 were excluded. The association test was conducted under the Cox’s proportional hazards model, which implies a positive effect size for a longer life. Transcriptome-wide association studies (TWAS) integrate GWAS and eQTLs to identify genes with a genetically regulated level of expression (eGenes) associated with human quantitative traits and/or diseases. In an attempt to implicate eGenes in the etiology of parental lifespan across multiple tissues, Timmers et al.^4^ used Summary-level Mendelian Randomization and Heterogeneity in Independent Instruments (HEIDI) tests from eQTL mapping in 48 tissues of the GTEx consortium. Although this approach helped identify the causal gene implicated in parental lifespan from the main GWAS signal, few if any new longevity genes were identified using TWAS. We used data from the Genotype-Tissue Expression Project (GTEx) resource (version 8) to perform TWAS on parental lifespan. GTEx is a large-scale multi-omic dataset where DNA and RNA were collected postmortem from 53 tissue samples from 635 donors. Tissues with less than 70 samples were removed to provide sufficient statistical power for eQTL discovery, resulting in a set of 48 tissues. Only non-sex-specific tissues (N=43) were analyzed. Alignment to the human reference genome hg28/GRCh38 was performed using STAR v2.6.1d, based on the GENCODE v30 annotation. RNA-seq expression outliers were excluded using a multidimensional extension of the statistic described by Wright et al.^48^ Samples with less than 10 million mapped reads were removed. For samples with replicates, replicate with the greatest number of reads were selected. Expression values were normalized between samples using TMM as implemented in edgeR^49^. For each gene, expression values were normalized across samples using an inverse normal transformation. Gene-level analyses were performed with S-PrediXcan^6^, which estimates cis-eQTL effect sizes with reference datasets. Resulting eQTL effects sizes were used to impute eGenes that were tested using summary statistics of the parental lifespan GWAS described above. S-PrediXcan enables TWAS without the need of individual-level data based on the MetaXcan framework. S-PrediXcan uses a parametric model (elastic net) that assumes a combination of LASSO and Ridge penalties on the eQTL effect sizes. eQTL prediction models were performed using elastic net, a regularized regression method, as implemented in the PredictDB pipeline^6,50^. We used SNPs with a minor allele frequency greater than 1% from European ancestry participants. Expression of protein coding, antisense, long intergenic non-coding and micro RNA was considered. The first three ancestry-based principal components, a set of covariates identified using the probabilistic estimation of expression residuals method^51^ with genotyping platform and sex were used as covariates.

### Assessment of genetic colocalization

Genetic colocalization was used to filter our LD-contaminated associations. A stringent Bayesian model is used to estimate the posterior probability of each eGene containing a single eQTL affecting both the eGene and the outcome(s) of interest. We used COLOC R package to estimate the probability of five hypotheses: H0 corresponds to no eQTL and no GWAS association, H1 and H2 correspond to eQTL association but no GWAS association or vice-versa, H3 corresponds to eQTL and GWAS associations but independent signals (weaker eQTL signal or GWAS hit) and H4 corresponds to shared eQTL and GWAS signal (the lead eQTL is also amongst the top GWAS hits)^52^. We kept eGenes with evidence of genetic colocalization (PP.H4>0.75). *Locuscompare* function from the *LocuscompareR* R package^9^ was used to depict the colocalization events. This software enables visualization of the strengths of eQTLs and outcomes associations by plotting p-values for each within a given genomic location, thereby contributing to distinguish candidates from false-positive genes.

### Tissue-specificity of gene expression and analysis of single-cell sequencing data of human livers

The tissue-specific gene expression metric (Tau) was obtained from all genes identified by TWAS. We used the formula from Yanai et al.^53^ to compare the level of gene expression across selected tissues based on RNA sequencing data from European ancestry donors from GTEx. All the genes with expression <1 RPKM were set as not expressed. The RNA-seq data were first log-transformed. After the normalization, a mean value from all replicates for each tissue separately was calculated. A Tau value closer to 1 indicates tissue-specificity while a Tau value closer to 0 indicates ubiquitous gene expression. We considered that eGenes had tissue-specific expression when their Tau statistic was ≥0.80. As the *HP* gene showed very high tissue specificity and since it was the only gene with a strong GWAS, TWAS and proteome-wide MR signal, we interrogated the web portal of the Human liver cell atlas recently made available by Aizarani et al.^8^. Briefly, this tool enables visualization of gene expression across clusters of liver cells that were obtained following analysis of over 10,000 unique liver cells obtained from 9 healthy donors. Clusters of cells included, B cells, Kupffer cells, NK, NKT and T cells, hepatocytes as well as a variety of endothelial cells and myofibroblasts. We visually inspected the cellular distribution of *HP* expression and compared it to that of *ASGR1* (asialoglycoprotein receptor 1), which has been described as a marker of hepatocytes^54^ for this analysis and *GAPDH*, a ubiquitously expressed gene.

### Associations of eProteins and eMetabolites with parental lifespan

We used GWAS summary statistics from the INTERVAL cohort^10^ to identify eProteins that could potentially be causally associated with parental lifespan. Relative concentrations of 3,622 plasma proteins or protein complexes were assayed using an aptamer-based multiplex protein assay (SOMAscan) in 3,301 participants from the INTERVAL study. A minimum of 4 instrumental variables (IVs) were constructed using cis-pQTLs in a 1Mb window, clumped using *plink 1.9* with a r^2^ < 0.1 (from 1000 genomes phase 3 European ancestry LD reference panel), a p-value <5e-8, and a physical distance threshold of 250 Kb. The MHC (6:28,477,797-33,448,354) and APOE regions were removed as well as ABO gene and immunoglobulins. Association of 279 circulating eProteins and parental lifespan was assessed with inverse variance weighted MR (IVW-MR) using *mr* function from *TwoSampleMR* package and a Bonferroni correction was applied (p-value = 1.79×10^−4^ (0.05/279 eProteins). The IVW-MR is comparable to performing a meta-analysis of each Wald ratio (the effect of the genetic instrument on eGenes divided by its effect on outcomes). Additional MR analyses were performed to evaluate heterogeneity (intercept p-value, from MR Egger^55^) and the presence of outliers. We used MR-PRESSO^56^, an outlier-robust method, to detect the presence of outliers (variants potentially causing pleiotropy and influencing causal estimates) and causal estimates were obtained before and after excluding outliers. We also used the contamination mixture method as this method was recently identified as a robust modelling method to identify causal relationships in the presence of potentially invalid genetic instruments^57^. The same MR framework was used to identify eMetabolites potentially associated with parental lifespan. We used GWAS summary on 123 metabolites from the study of Kettunen et al.^11^ In this study, 123 blood lipids and metabolites were measured in 24,925 individuals from 10 European cohorts using high-throughput nuclear magnetic resonance spectroscopy. Metabolites measured by NMR represent a broad molecular signature of systemic metabolism and includes metabolites from multiple metabolic pathways (lipoprotein lipids and subclasses, fatty acids as well as amino acids, glycolysis precursors, etc.). IVs for eMetabolites were constructed using independent genetic variants clumped using *plink 1.9* with a r^2^ < 0.1, a p-value <5e-8 and a physical distance threshold of 250 Kb. Association of 115 circulating eMetabolites and parental lifespan was assessed with IVW-MR using *mr* function from *TwoSampleMR* package and a Bonferroni correction was applied (p-value = 4.35×10^−4^ (0.05/115 eMetabolite). MR-PRESSO, MR-Egger and contamination mixture methods were also used as described above.

### Association of eGenes, eProteins, eMetabolites with disease traits

For MR analysis, cis-eQTL were identified using QTLtools^58^ with the first three principal components, a set of covariates identified using the probabilistic estimation of expression residuals method^51^ with genotyping platform and sex as covariates. Variants were included if they had a minor allele frequency ≥ 1%. Missing genotypes were imputed as the mean of the other participants’ genotypes. The cis-window size was set to 1Mb. We used the *get_r_from_pn* and *get_r_from_lor* functions from the *TwoSampleMR* package to calculate variance of each eQTL for quantitative and binary data respectively. SNPs that explain more of the variance in the outcome compared to the exposure was not included as instrumental variables (IV). Removing eQTLs that show evidence of reverse causality is of particular importance in the setting of human longevity studies as aging might influence genes in a tissue-specific manner^59^. IVs were clumped using plink 1.9 with a r^2^ < 0.1, a p-value < 0.01 and a physical distance threshold of 250 Kb. If an eGene was significant in multiple tissues, the tissue that provided the strongest eGenes-parental lifespan estimate from S-PrediXcan was prioritized. Instruments for eProteins and eMetabolites were the same as those used to determine their association with parental lifespan. A detailed description of the GWAS summary statistics and cohorts used for this analysis is presented in Supplementary Table 5. To increase statistical power in some of the studies, we performed fixed-effects GWAS meta-analysis of traits of the UK Biobank and FinnGen cohorts (selected traits described below) using the METAL software^60^. Altogether, we tested the association between 48 instruments (31 eGenes, 9 eProteins, 3 eMetabolites and 5 SNPs) and 40 age-associated chronic diseases with at least 1000 cases. The statistical significance threshold was set accordingly (0.05/(40×48) = 2.60×10^−5^). SNPs, eGenes or eProteins in and around the major histocompatibility complex region were not included in this analysis.

### Multi-trait colocalization

We investigated shared genetic etiology at chromosome 15 within 1 Mb of the *HP* gene between liver expression of the *HP* gene (from GTEx^61^), plasma Hp levels (from INTERVAL^10^), plasma LDL-C levels (from GLGC^62^) and parental lifespan (from UK Biobank and LifeGen^4^) using default settings of the *hyprcoloc* R package. HyPrColoc is a deterministic Bayesian algorithm using GWAS summary statistics that can detect colocalization across vast numbers of traits simultaneously^21^. Posterior probabilities of colocalization heatmap was performed with the *sensitivity.plot* function from the *hyprcoloc* R package with default settings (regional and alignment thresholds: 0.6, 0.7, 0.8 and 0.9, prior probabilities of colocalization: 0.98, 0.99 and 0.995). Regional association plot was obtained from the *stack_assoc_plot* function from the *gassocplot* R package and the 1000G phase 3 LD reference panel (European ancestry).

### Resources used to identify drug candidates for human healthspan extension

We used the Open Targets Platform to determine the potential therapeutic areas of the eGenes and eProteins associated with parental lifespan and to identify approved therapies that may target these eGenes or eProteins. Briefly, the Open Targets Platform developed a user-friendly web interface (available at: https://www.targetvalidation.org/) that enables the search of therapeutic areas, drug targets, pathways, gene ontology, and tractability information simultaneous for gene lists prepared by the user. This information is made available by the platform’s integration of evidence from genetics, genomics, transcriptomics, drugs, animal models and scientific literature to score and rank target-diseases associations for drug target identification. We also used the DrugBank encyclopedia to identify potential drug repositioning opportunities and drug targets that may interact with eGenes and eProteins identified in the course of this investigation. DrugBank is a publicly available resource with drug and drug-target information on over 13,000 drug entries available at: https://www.drugbank.ca/. The Drug Gene Interaction database is an open source resource than enables the query of genes of interest with respect to known drug-gene interactions and potential druggability, it is available at: http://www.dgidb.org/ ^2^. We also queried eGenes and eProteins in the GeneMANIA prediction server to gain new information on pathways and genetic interactions mostly connected with the eGenes and eProteins^63^. Briefly, GeneMANIA integrates data from thousands of genomics and proteomics datasets such as the Gene Expression Omnibus (GEO), BioGRID, IRefindex, I2D as well as organisms-specific functional genomics datasets to enable users to query list of genes in a user-friendly web portal. GeneMANIA algorithms use patterns of gene co-annotations in the Gene Ontology (GO) biological functions hierarchy, which are weighted to find closely connected genes and generates networks of gene co-expression, proteins with physical interactions, shared domains, etc.

### Variation in PCSK9 and parental lifespan using individual participant data in the UK Biobank

We selected independent SNPs (*r*^*2*^<0.10) at the *PCSK9* locus (within 100 Kb of the gene) associated with LDL-C levels at a genome-wide level of significance in the Global Lipids Genetics Consortium (GLGC)^62^. This approach yielded 11 SNPs independently associated with LDL-C levels. We constructed a weighted genetic score (GS) of SNPs at the *PCSK9* locus scaled to a 1 mmol/L reduction in LDL-C levels. The associations between the GS and parental lifespan in the UK Biobank was tested using logistic regression adjusted for age, sex and the first 10 ancestry-based principal components using R (version 3.5.1). We used the definition of Pilling et al.^64^ to assess high parental lifespan in participants of the UK Biobank. Only participants between 55 and 69 years were included. Participants who were adopted, had missing information on age of parents’ death or who had parents who died at a young age (<46 for the father and <57 for the mother) were excluded from these analyses. Parents were separated into three categories: long-lived (father still alive and older than 90 or father’s age of death ≥90 and mother still alive and older than 93 or mother’s age of death ≥93), medium lived (age of death ≥66 and <89 for the father and ≥73 and <92 for the mother) and short lived (age of death ≥46 and <65 for the father and ≥57 and <72 for the mother). We defined high parental lifespan as at least one long-lived parent (i.e. long/long or long/medium). The control group included participants with parents considered as short- or medium-lived (i.e. short/short, short/medium or medium/medium). Participants discordant for mothers’ and fathers’ age of death (one long-lived parent and one short-lived parent) were also excluded from the present analyses. These analyses were conducted under UK Biobank data application number 25205.

### Phenome-wide mendelian randomization studies in the UK Biobank and FinnGen cohorts

We obtained GWAS summary statistics on 1403 disease-specific binary traits in 408,961 participants of the UK Biobank to identify diseases that may be influenced by the longevity loci that were identified in previous steps of this investigation^13^. In this study, a method called SAIGE (Scalable and Accurate Implementation of Generalized Mixed Models), which is based on generalized mixed models was developed to control for case-control imbalance, sample relatedness and population structure. A previously published scheme was used to defined disease-specific binary traits by combining International Classification of Diseases (ICD)-9 codes into hierarchical “PheCodes”. UK Biobank participants were assigned a PheCode if they had one or more of the PheCode-specific ICD codes. GWAS was performed using 28 million genetic markers directly genotyped or imputed by the Haplotype Reference Consortium (HRC) panel with SAIGE, adjusting for sex and birth year. Sex-specific outcomes and outcomes with a case: control ratio <1:1000 were excluded leaving 791 traits for PheWAS. We considered associations that had a p-value <6.3×10^−5^ (0.05/791 traits) to be statistically significant. SAIGE was also used to obtain GWAS summary statistics in the FinnGen cohorts. GWAS was performed using over 16 million genetic markers genotyped with the Illumina or Affymetrix arrays or imputed using the population specific SISu v3 reference panel. Variables included in the models were sex, age, the 10-main ancestry-based principal components and genotyping batch. This analysis was performed using publicly available GWAS summary statistics from the FinnGen data freeze 2 (January 14, 2020). Sex-specific outcomes and outcomes with <400 cases were excluded leaving 809 traits for PheWAS. We considered associations that had a p-value <6.2×10^−5^ (0.05/809 traits) to be statistically significant. In both datasets (UK Biobank and FinnGen), we used IVW-MR to determine the association between genetic instruments for eGenes, eProteins, eMetabolites or single SNPs without TWAS (*APOE, LPA, LDLR, MAGI3* and 9p21 region) these disease-specific binary traits. We used genetic instruments mentioned above while keeping the SNPs in common between the compared datasets.

### Data availability

The summary statistics of the GWAS meta-analysis of parental lifespan (Timmers et al.) are available for download at https://datashare.is.ed.ac.uk/handle/10283/3209.

GTEx data is available to download at https://gtexportal.org/home/datasets. The data used for the analyses described in this manuscript were obtained from dbGaP, accession number <u>phs000424.vN.pN</u>.

The weblinks to the GWAS summary statistics files of the 40 chronic diseases are available in Supplementary Table 5. The GWAS summary statistics for >1400 binary phenotypes in the UK Biobank by SAIGE are available to download at https://www.leelabsg.org/resources. The GWAS summary statistics for >1100 binary phenotypes in the FinnGen cohorts by SAIGE are available to download at https://www.finngen.fi/en/access_results. GWAS summary statistics for the proteins of the INTERVAL cohort are available for download at: https://www.phpc.cam.ac.uk/ceu/proteins/

GWAS summary statistics for lipoprotein metabolomics parameters, from Kettunen et al. are available for download at: http://www.computationalmedicine.fi/data#NMR_GWAS. The MetaXcan software is available to download at https://github.com/hakyimlab/MetaXcan.

*TwoSampleMR* R package is available at https://mrcieu.github.io/TwoSampleMR/. *Coloc* R package is available at https://github.com/chr1swallace/coloc/. *Hyprcoloc* R package is available at https://github.com/jrs95/hyprcoloc/. *Gassocplot* R package is available at https://github.com/jrs95/gassocplot. *LocusCompareR* R package is available at https://github.com/boxiangliu/locuscomparer. The Human liver cell atlas of Aizarani et al.^8^ is available at http://human-liver-cell-atlas.ie-freiburg.mpg.de/.

## Notes

### Competing Interest Statement

The authors have declared no competing interest.

### Funding Statement

NP holds a doctoral research award from the Fonds de recherche du Quebec: Sante. (FRQS). BJA and ST hold junior scholar awards from the FRQS. PM holds a FRQS Research Chair on the Pathobiology of Calcific Aortic Valve Disease. YB holds a Canada Research Chair in Genomics of Heart and Lung Diseases.

### Author Declarations

Because the data included in this manuscript was obtained from publicly available sources, no ethical approval was obtained for this specific project. Analyses performed with the UK Biobank dataset were conducted under UK Biobank data application number 25205. This project was approved by the IRB of the Quebec Heart and Lung Institute.

## REFERENCES

1 Melzer, D., Pilling, L. C. & Ferrucci, L. The genetics of human ageing. Nat Rev Genet 21, 88–101, doi:10.1038/s41576-019-0183-6 (2020).

2 Cotto, K. C. et al.. DGIdb 3.0: a redesign and expansion of the drug-gene interaction database. Nucleic Acids Res 46, D1068–D1073, doi:10.1093/nar/gkx1143 (2018).

3 Deelen, J. et al.. A meta-analysis of genome-wide association studies identifies multiple longevity genes. Nat Commun 10, 3669, doi:10.1038/s41467-019-11558-2 (2019).

4 Timmers, P. R. et al.. Genomics of 1 million parent lifespans implicates novel pathways and common diseases and distinguishes survival chances. Elife 8, doi:10.7554/eLife.39856 (2019).

5 Hemani, G. et al.. The MR-Base platform supports systematic causal inference across the human phenome. Elife 7, doi:10.7554/eLife.34408 (2018).

6 Barbeira, A. N. et al.. Exploring the phenotypic consequences of tissue specific gene expression variation inferred from GWAS summary statistics. Nat Commun 9, 1825, doi:10.1038/s41467-018-03621-1 (2018).

7 Kryuchkova-Mostacci, N. & Robinson-Rechavi, M. A benchmark of gene expression tissue-specificity metrics. Brief Bioinform 18, 205–214, doi:10.1093/bib/bbw008 (2017).

8 Aizarani, N. et al.. A human liver cell atlas reveals heterogeneity and epithelial progenitors. Nature 572, 199–204, doi:10.1038/s41586-019-1373-2 (2019).

9 Liu, B., Gloudemans, M. J., Rao, A. S., Ingelsson, E. & Montgomery, S. B. Abundant associations with gene expression complicate GWAS follow-up. Nat Genet 51, 768–769, doi:10.1038/s41588-019-0404-0 (2019).

10 Sun, B. B. et al.. Genomic atlas of the human plasma proteome. Nature 558, 73–79, doi:10.1038/s41586-018-0175-2 (2018).

11 Kettunen, J. et al.. Genome-wide study for circulating metabolites identifies 62 loci and reveals novel systemic effects of LPA. Nat Commun 7, 11122, doi:10.1038/ncomms11122 (2016).

12 van der Harst, P. & Verweij, N. Identification of 64 Novel Genetic Loci Provides an Expanded View on the Genetic Architecture of Coronary Artery Disease. Circ Res 122, 433–443, doi:10.1161/CIRCRESAHA.117.312086 (2018).

13 Zhou, W. et al.. Efficiently controlling for case-control imbalance and sample relatedness in large-scale genetic association studies. Nat Genet 50, 1335–1341, doi:10.1038/s41588-018-0184-y (2018).

14 Malik, R. et al.. Multiancestry genome-wide association study of 520,000 subjects identifies 32 loci associated with stroke and stroke subtypes. Nat Genet 50, 524–537, doi:10.1038/s41588-018-0058-3 (2018).

15 Shah, S. et al.. Genome-wide association and Mendelian randomisation analysis provide insights into the pathogenesis of heart failure. Nat Commun 11, 163, doi:10.1038/s41467-019-13690-5 (2020).

16 Watanabe, K. et al.. A global overview of pleiotropy and genetic architecture in complex traits. Nat Genet 51, 1339–1348, doi:10.1038/s41588-019-0481-0 (2019).

17 Mahajan, A. et al.. Fine-mapping type 2 diabetes loci to single-variant resolution using high-density imputation and islet-specific epigenome maps. Nat Genet 50, 1505–1513, doi:10.1038/s41588-018-0241-6 (2018).

18 Wuttke, M. et al.. A catalog of genetic loci associated with kidney function from analyses of a million individuals. Nat Genet 51, 957–972, doi:10.1038/s41588-019-0407-x (2019).

19 Namjou, B. et al.. GWAS and enrichment analyses of non-alcoholic fatty liver disease identify new trait-associated genes and pathways across eMERGE Network. BMC Med 17, 135, doi:10.1186/s12916-019-1364-z (2019).

20 Jansen, I. E. et al.. Genome-wide meta-analysis identifies new loci and functional pathways influencing Alzheimer’s disease risk. Nat Genet 51, 404–413, doi:10.1038/s41588-018-0311-9 (2019).

21 Foley, C. N. et al.. A fast and efficient colocalization algorithm for identifying shared genetic risk factors across multiple traits. bioRxiv, 592238, doi:10.1101/592238 (2019).

22 Carvalho-Silva, D. et al.. Open Targets Platform: new developments and updates two years on. Nucleic Acids Res 47, D1056–D1065, doi:10.1093/nar/gky1133 (2019).

23 Wishart, D. S. et al.. DrugBank 5.0: a major update to the DrugBank database for 2018. Nucleic Acids Res 46, D1074–D1082, doi:10.1093/nar/gkx1037 (2018).

24 Franz, M. et al.. GeneMANIA update 2018. Nucleic Acids Res 46, W60–W64, doi:10.1093/nar/gky311 (2018).

25 Rashid, S. et al.. Decreased plasma cholesterol and hypersensitivity to statins in mice lacking Pcsk9. Proc Natl Acad Sci U S A 102, 5374–5379, doi:10.1073/pnas.0501652102 (2005).

26 Kosenko, T., Golder, M., Leblond, G., Weng, W. & Lagace, T. A. Low density lipoprotein binds to proprotein convertase subtilisin/kexin type-9 (PCSK9) in human plasma and inhibits PCSK9-mediated low density lipoprotein receptor degradation. J Biol Chem 288, 8279–8288, doi:10.1074/jbc.M112.421370 (2013).

27 Tavori, H. et al.. PCSK9 Association With Lipoprotein(a). Circ Res 119, 29–35, doi:10.1161/CIRCRESAHA.116.308811 (2016).

28 Benjannet, S., Rhainds, D., Hamelin, J., Nassoury, N. & Seidah, N. G. The proprotein convertase (PC) PCSK9 is inactivated by furin and/or PC5/6A: functional consequences of natural mutations and post-translational modifications. J Biol Chem 281, 30561–30572, doi:10.1074/jbc.M606495200 (2006).

29 Chignon, A. et al.. Single-cell expression and Mendelian randomization analyses identify blood genes associated with lifespan and chronic diseases. Commun Biol 3, 206, doi:10.1038/s42003-020-0937-x (2020).

30 Pedersen, T. R. The Success Story of LDL Cholesterol Lowering. Circ Res 118, 721–731, doi:10.1161/CIRCRESAHA.115.306297 (2016).

31 Collins, R. et al.. Interpretation of the evidence for the efficacy and safety of statin therapy. Lancet 388, 2532–2561, doi:10.1016/S0140-6736(16)31357-5 (2016).

32 Defesche, J. C. et al.. Familial hypercholesterolaemia. Nat Rev Dis Primers 3, 17093, doi:10.1038/nrdp.2017.93 (2017).

33 Benn, M., Nordestgaard, B. G., Frikke-Schmidt, R. & Tybjaerg-Hansen, A. Low LDL cholesterol, PCSK9 and HMGCR genetic variation, and risk of Alzheimer’s disease and Parkinson’s disease: Mendelian randomisation study. BMJ 357, j1648, doi:10.1136/bmj.j1648 (2017).

34 Brumpton, B. M. et al.. Variation in Serum PCSK9 (Proprotein Convertase Subtilisin/Kexin Type 9), Cardiovascular Disease Risk, and an Investigation of Potential Unanticipated Effects of PCSK9 Inhibition. Circ Genom Precis Med 12, e002335, doi:10.1161/CIRCGEN.118.002335 (2019).

35 Tsimikas, S. et al.. NHLBI Working Group Recommendations to Reduce Lipoprotein(a)- Mediated Risk of Cardiovascular Disease and Aortic Stenosis. J Am Coll Cardiol 71, 177–192, doi:10.1016/j.jacc.2017.11.014 (2018).

36 Arsenault, B. J. et al.. Association of Long-term Exposure to Elevated Lipoprotein(a) Levels With Parental Life Span, Chronic Disease-Free Survival, and Mortality Risk: A Mendelian Randomization Analysis. JAMA Netw Open 3, e200129, doi:10.1001/jamanetworkopen.2020.0129 (2020).

37 Musunuru, K. et al.. From noncoding variant to phenotype via SORT1 at the 1p13 cholesterol locus. Nature 466, 714–719, doi:10.1038/nature09266 (2010).

38 Kjolby, M. et al.. Sort1, encoded by the cardiovascular risk locus 1p13.3, is a regulator of hepatic lipoprotein export. Cell Metab 12, 213–223, doi:10.1016/j.cmet.2010.08.006 (2010).

39 Miura, Y. et al.. Proteomic analysis of plasma proteins in Japanese semisuper centenarians. Exp Gerontol 46, 81–85, doi:10.1016/j.exger.2010.10.002 (2011).

40 Boettger, L. M. et al.. Recurring exon deletions in the HP (haptoglobin) gene contribute to lower blood cholesterol levels. Nat Genet 48, 359–366, doi:10.1038/ng.3510 (2016).

41 Spagnuolo, M. S. et al.. Haptoglobin interacts with apolipoprotein E and beta-amyloid and influences their crosstalk. ACS Chem Neurosci 5, 837–847, doi:10.1021/cn500099f (2014).

42 Campisi, J. et al.. From discoveries in ageing research to therapeutics for healthy ageing. Nature 571, 183–192, doi:10.1038/s41586-019-1365-2 (2019).

43 Rosa, M. et al.. A Mendelian randomization study of IL6 signaling in cardiovascular diseases, immune-related disorders and longevity. NPJ Genom Med 4, 23, doi:10.1038/s41525-019-0097-4 (2019).

44 Ridker, P. M. et al.. Relationship of C-reactive protein reduction to cardiovascular event reduction following treatment with canakinumab: a secondary analysis from the CANTOS randomised controlled trial. Lancet 391, 319–328, doi:10.1016/S0140-6736(17)32814-3 (2018).

45 Prieto, M. L. et al.. Leveraging electronic health records to study pleiotropic effects on bipolar disorder and medical comorbidities. Transl Psychiatry 6, e870, doi:10.1038/tp.2016.138 (2016).

46 Myocardial Infarction, G. et al. Coding Variation in ANGPTL4, LPL, and SVEP1 and the Risk of Coronary Disease. N Engl J Med 374, 1134–1144, doi:10.1056/NEJMoa1507652 (2016).

47 Yashin, A. I., Wu, D., Arbeev, K. G. & Ukraintseva, S. V. Joint influence of small-effect genetic variants on human longevity. Aging (Albany NY) 2, 612–620, doi:10.18632/aging.100191 (2010).

48 Wright, F. A. et al.. Heritability and genomics of gene expression in peripheral blood. Nat Genet 46, 430–437, doi:10.1038/ng.2951 (2014).

49 Robinson, M. D. & Oshlack, A. A scaling normalization method for differential expression analysis of RNA-seq data. Genome Biol 11, R25, doi:10.1186/gb-2010-11-3-r25 (2010).

50 Gamazon, E. R. et al.. A gene-based association method for mapping traits using reference transcriptome data. Nat Genet 47, 1091–1098, doi:10.1038/ng.3367 (2015).

51 Stegle, O., Parts, L., Piipari, M., Winn, J. & Durbin, R. Using probabilistic estimation of expression residuals (PEER) to obtain increased power and interpretability of gene expression analyses. Nat Protoc 7, 500–507, doi:10.1038/nprot.2011.457 (2012).

52 Giambartolomei, C. et al.. Bayesian test for colocalisation between pairs of genetic association studies using summary statistics. PLoS Genet 10, e1004383, doi:10.1371/journal.pgen.1004383 (2014).

53 Yanai, I. et al.. Genome-wide midrange transcription profiles reveal expression level relationships in human tissue specification. Bioinformatics 21, 650–659, doi:10.1093/bioinformatics/bti042 (2005).

54 Peters, D. T. et al.. Asialoglycoprotein receptor 1 is a specific cell-surface marker for isolating hepatocytes derived from human pluripotent stem cells. Development 143, 1475–1481, doi:10.1242/dev.132209 (2016).

55 Bowden, J. et al.. A framework for the investigation of pleiotropy in two-sample summary data Mendelian randomization. Stat Med 36, 1783–1802, doi:10.1002/sim.7221 (2017).

56 Verbanck, M., Chen, C. Y., Neale, B. & Do, R. Detection of widespread horizontal pleiotropy in causal relationships inferred from Mendelian randomization between complex traits and diseases. Nat Genet 50, 693–698, doi:10.1038/s41588-018-0099-7 (2018).

57 Burgess, S., Foley, C. N., Allara, E., Staley, J. R. & Howson, J. M. M. A robust and efficient method for Mendelian randomization with hundreds of genetic variants. Nat Commun 11, 376, doi:10.1038/s41467-019-14156-4 (2020).

58 Delaneau, O. et al.. A complete tool set for molecular QTL discovery and analysis. Nat Commun 8, 15452, doi:10.1038/ncomms15452 (2017).

59 Yang, J. et al.. Synchronized age-related gene expression changes across multiple tissues in human and the link to complex diseases. Sci Rep 5, 15145, doi:10.1038/srep15145 (2015).

60 Willer, C. J., Li, Y. & Abecasis, G. R. METAL: fast and efficient meta-analysis of genomewide association scans. Bioinformatics 26, 2190–2191, doi:10.1093/bioinformatics/btq340 (2010).

61 Consortium, G. T. The Genotype-Tissue Expression (GTEx) project. Nat Genet 45, 580–585, doi:10.1038/ng.2653 (2013).

62 Willer, C. J. et al.. Discovery and refinement of loci associated with lipid levels. Nat Genet 45, 1274–1283, doi:10.1038/ng.2797 (2013).

63 Warde-Farley, D. et al.. The GeneMANIA prediction server: biological network integration for gene prioritization and predicting gene function. Nucleic Acids Res 38, W214–220, doi:10.1093/nar/gkq537 (2010).

64 Pilling, L. C. et al.. Human longevity: 25 genetic loci associated in 389,166 UK biobank participants. Aging (Albany NY) 9, 2504–2520, doi:10.18632/aging.101334 (2017).

